# Predicting Clinical Endpoints and Visual Changes with Quality-Weighted Tissue-based Renal Histological Features

**DOI:** 10.1101/2022.03.30.22269826

**Authors:** Ka Ho Tam, Maria F. Soares, Jesper Kers, Edward J. Sharples, Rutger J. Ploeg, Maria Kaisar, Jens Rittscher

## Abstract

Two common obstacles limiting the performance of data-driven algorithms in digital histopathology classification tasks are the lack of expert annotations and the narrow diversity of datasets. Multi-instance learning (MIL) can be used to address the former challenge for the analysis of whole slide images (WSI) but performance is often inferior to full supervision. We show that the inclusion of weak annotations can significantly enhance the effectiveness of MIL while keeping the approach scalable.

An analysis framework was developed to process periodic acid-Schiff (PAS) and Sirius Red (SR) slides of renal biopsies. The workflow segments tissues into coarse tissue classes. Handcrafted and deep features were extracted from these tissues and combined using a soft attention model to predict several slide-level labels: delayed graft function (DGF), acute tubular injury (ATI), and Remuzzi grade components. A tissue segmentation quality metric was also developed to reduce the adverse impact of poorly segmented instances. The soft attention model was trained using 5-fold cross-validation on a mixed dataset and tested on the QUOD dataset containing n=373 PAS and n=195 SR biopsies.

The average ROC-AUC over different prediction tasks was found to be 0.598±0.011, significantly higher than using only ResNet50 (0.545±0.012), only handcrafted features (0.542±0.011), and the baseline (0.532±0.012) of state-of-the-art performance. Weighting tissues by segmentation quality in conjunction with soft attention has led to further improvement (*AUC* = 0.618 ± 0.010). Using an intuitive visualisation scheme, we show that our approach may also be used to support clinical decision making as it allows pinpointing individual tissues relevant to the predictions.

**2020 MSC:** 68T07, 92C50

## 1. Introduction

Computational pathology can assist pathologists by providing an automated second opinion on their assessment. Moreover, it may help us to better understand the mechanisms of organ injury by detecting and quantifying subtle histological changes in biopsies. While these models can help us improve the discriminative power of assessment tasks with performance unrivaled by classical image processing algorithms, there are several challenges limiting their applicability. Firstly, training neural networks often requires large amounts of labelled data. However, most datasets contain no more than several hundred slides. In our setting, samples with known outcomes are biased due to pretransplantation screening (either based on the patient’s clinical information or histology). The only available data are “hard examples” that have either been missed by pathologists or are plagued by factors not guaranteed to be visible in biopsies. There is also a “bootstrap” problem - while a severe shortage of pathologists is a primary motivation to expedite the development of an automated tool, this shortage limits the speed and scale in which labelled data can be procured.

To date, most deep-learning based computational pathology platforms are designed to predict or assess only a bespoke set of narrowly defined clinical outcomes or visual changes (collectively known as slide-level labels). Most existing work (Yi et al., 2022; Hermsen et al., 2019; Marsh et al., 2018; Davis et al., 2021; Kers et al., 2022) is limited to fully-supervised learning, which requires labelling large number of tissue compartments or rectangular tiles as either normal or diseased. To adapt these platforms for a different diagnosis would require additional time from pathologists to go through the entire dataset, adding further to the project’s investment. Regrettably, the expert-time cost of fully-supervised learning is prohibitive and has been a major reason for the limited number of publications applied to renal histology.

To address the lack of local expert annotations, a number of multi-instance learning approaches (Lu et al., 2021; Iizuka et al., 2020; Campanella et al., 2019; Li et al., 2021) have been developed to train classification tasks using only slide-level labels. However, available multi-instance learning models typically need to be trained on either large datasets, or on slides with plenty of tissue area with good diagnostic quality. In our setting where many slides contain sub-optimal tissue areas, we find existing approaches fail to deliver acceptable classification performance.

Furthermore, models trained under multi-instance learning tend to have limited diagnostic transparency - features are often extracted from rectangular tiles which are inconsistent with the anatomical, irregular tissue compartments within the biopsies. It is common for different functional tissue structures to vary in size by several orders of magnitude (eg. cell nuclei vs arteries). This may be partially addressed by using tiles from several magnifications (Li et al., 2021), but this solution could make visualisation considerably more challenging.

Any histology analysis framework also needs to be robust to artifacts. This is particularly true in our application for two reasons. Firstly, in needle biopsies, a large proportion of tissue resides close to the edge and is often distorted or truncated. Exclusion of these tissues is not always possible as biopsies are often narrow and have limited material available. Secondly, decisions of transplantation are time-sensitive hence the long-term aim is to eventually read histology from frozen biopsies which are often plagued with artifacts. There is a lack of definitive attempts to reduce the impact of artifacts. Treatment of artifacts is often either not mentioned or is excluded manually in most experiments. To our knowledge, the most common approach to tackle artifacts include explicitly labelling of artifacts and aggressive data augmentation (Marsh et al., 2018; Davis et al., 2021). However, these approaches would only work on objects that resemble the training data.

The goal of this pilot study is to show the feasibility of a flexible yet scalable platform for providing quantitative insight into visual associations to transplant dysfunction using renal biopsies stained with periodic acid-Schiff (PAS) and Sirius Red (SR) while addressing the aforementioned issues. Our workflow extracts a number of histologically relevant visual features from tissues and was developed with minimal laborious labelling by expert pathologists. We used these visual features in combination with convolutional neural network (CNN) features to predict several slide-level labels such as Delayed Graft Function (DGF) - defined as patients who need dialysis within the first week after transplantation (Yarlagadda et al., 2008), Acute Tubular Injury (ATI), and Remuzzi Scores (Remuzzi et al., 2006). We compared the predictive performance of our framework with features based on tissue compartments with a standard workflow that relies only on CNN-derived features and on rectangular tiles and showed that the proposed workflow produces consistently higher area-under-the-curve (AUC) in the models’ receiver operator characteristics (ROC) and Precision-Recall (PR) curves.

Furthermore, we developed a visualisation scheme that works specifically for our proposed workflow. Compared to prior work (Lu et al., 2021; Iizuka et al., 2020; Campanella et al., 2019), using tissue-derived features enabled us to pinpoint a diagnosis to specific tissues irrespective of their size and shape, enabling the potential of transparent diagnosis and visualisation.

Finally, through our experiments, we also find that tissue quality and quantity may play a significant role in the predictability of a slide. By incorporating a metric derived from Bayesian Neural Networks (BNN) describing tissue quality similar to Tam et al. (Tam et al., 2020) derived from an ensemble of CNN models, we are able to consistently improve the quality of predictions measured by AUC.

## 2. Method

We propose a computational framework to extract visual histopathological features from different functional components of the biopsy. A schematic of the workflow is shown in Figure 1. In a first step we identify specific tissue compartments (Figure 1(a)). Details of the segmentation algorithm are discussed in Section 2.2. Subsequently, we extract a set of tissue compartment specific features (Figure 1(b)) which are described in Section 2.3).

**Figure 1:**
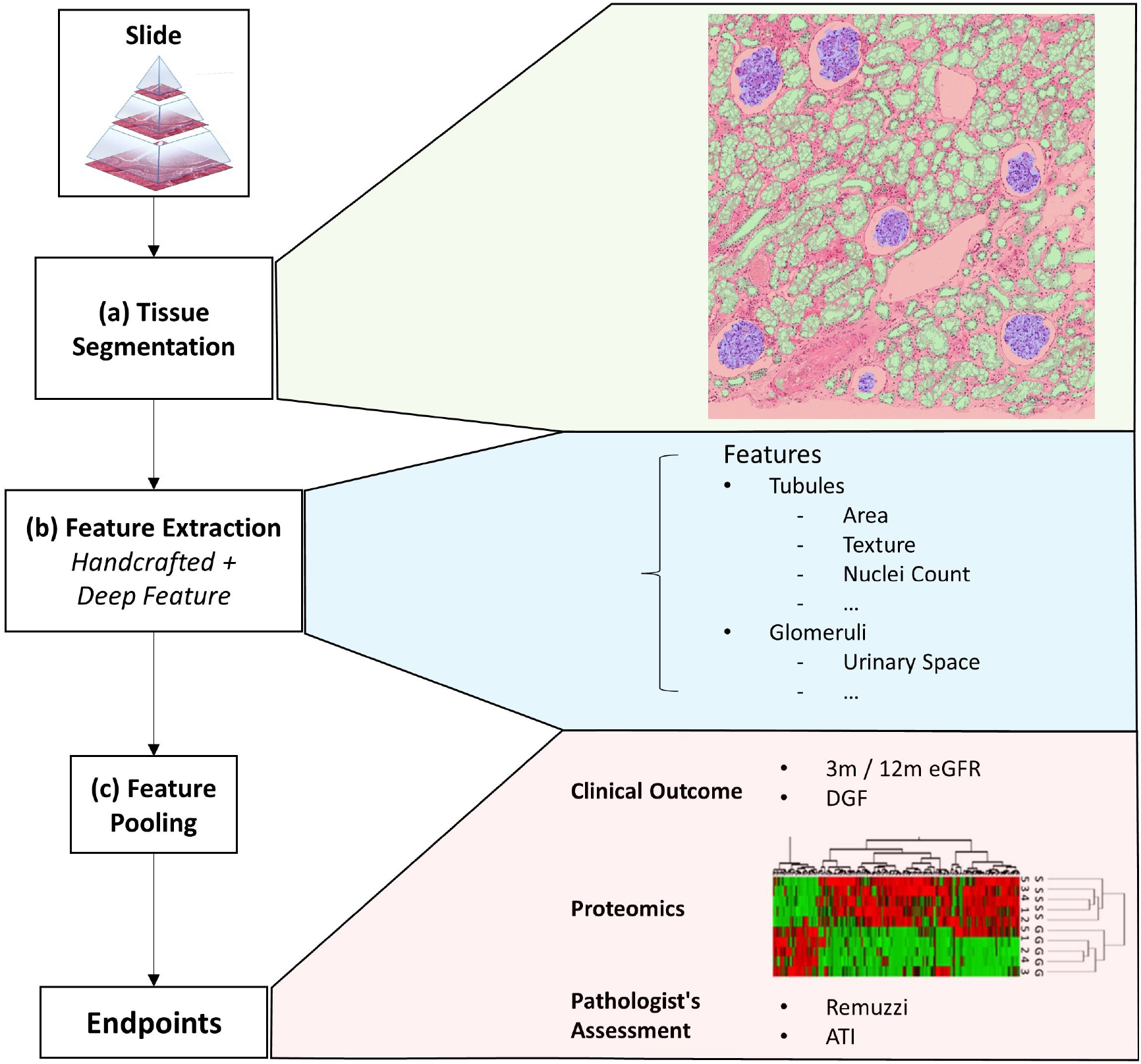
Overview of the framework. This figure shows an overview of the proposed quantitative analysis framework. (a) Tissue segmentation returns the instance outline of three different tissue types and cell nuclei. (b) Feature extraction returns a mixture of handcrafted and deep features iterated over each tissue. (c) Finally, features from a variable number of tissues are pooled together with soft attention to form a single vectorial description of a slide to predict the slide’s label. The slide label could either be clinical endpoints, assessment results given by pathologists, or other biomarkers.

Finally, we combine combine instance level features into a fixed-length description of the whole slide (Figure 1(c)). We evaluate different approaches to combining these features. The most trivial method is to simply perform an average/max pooling from the feature values of all the tissues. However, if the segmented tissues were only coarsely categorised, a large portion may be irrelevant for diagnosis. Pooling features from different tissues irrespective of their histopathological importance could lead to erroneous predictions that lack transparency.

Multi-instance learning (MIL) (Campanella et al., 2019; Maron and Lozano-Pérez, 1998; Ilse et al., 2018) is another approach commonly applied to histology analysis for making slide level (bag) predictions from a variable number of instances. A slide is classified as positive if at least one positive instance is detected. Implementation of the original MIL algorithm involves predicting a probability value for each instance and then converting these instance probabilities into a bag-level value using max-pooling. However, this approach has several limitations that make it unsuitable for predicting kidney function.

Firstly, the use of max-pooling means that the method is particularly sensitive to noise. In our slides, we often have artifacts or tissues with morphology not previously seen during training. There is an inherent risk that the presence of these artifacts could sway the predictions as the classifiers have not been trained on embeddings beyond the original data. Secondly, although we are formulating our problem as a classification task, kidney function and the grading of slides have an inherent progressive nature. Standard MIL is not well adapted to handling multi-class classification problems and it gives predictions that lack symmetry between the positives and negatives.

To partially mitigate the aforementioned limitations, we implement a soft attention mechanism. As a result we can use attention-weighted averages of the instances to make predictions as shown in the schematic in Figure 2. The model consists of multiple stages: Firstly, it converts each instance’s features into a permutation-invariant embedding. From these embeddings, a gated attention mechanism (Ilse et al., 2018) assigns weights to each instance depending on their relative importance for the bag-level predictions. The reason a gated mechanism is used is to enhance the non-linearity of *tanh* when the function’s input values are small. Because soft attention is learned, theoretically, it should be capable of rejecting instances not relevant for the assigned bag-level prediction task. The fractional contribution *a*_*k*_ of an instance *k* to the final prediction is:

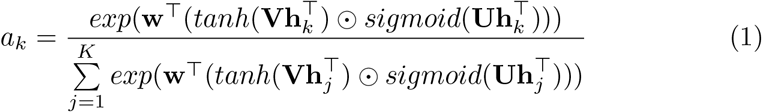

Where **h** is an embedding derived from the feature vector of one instance, **w, U, V** are learnable weights in our neural network.

**Figure 2:**
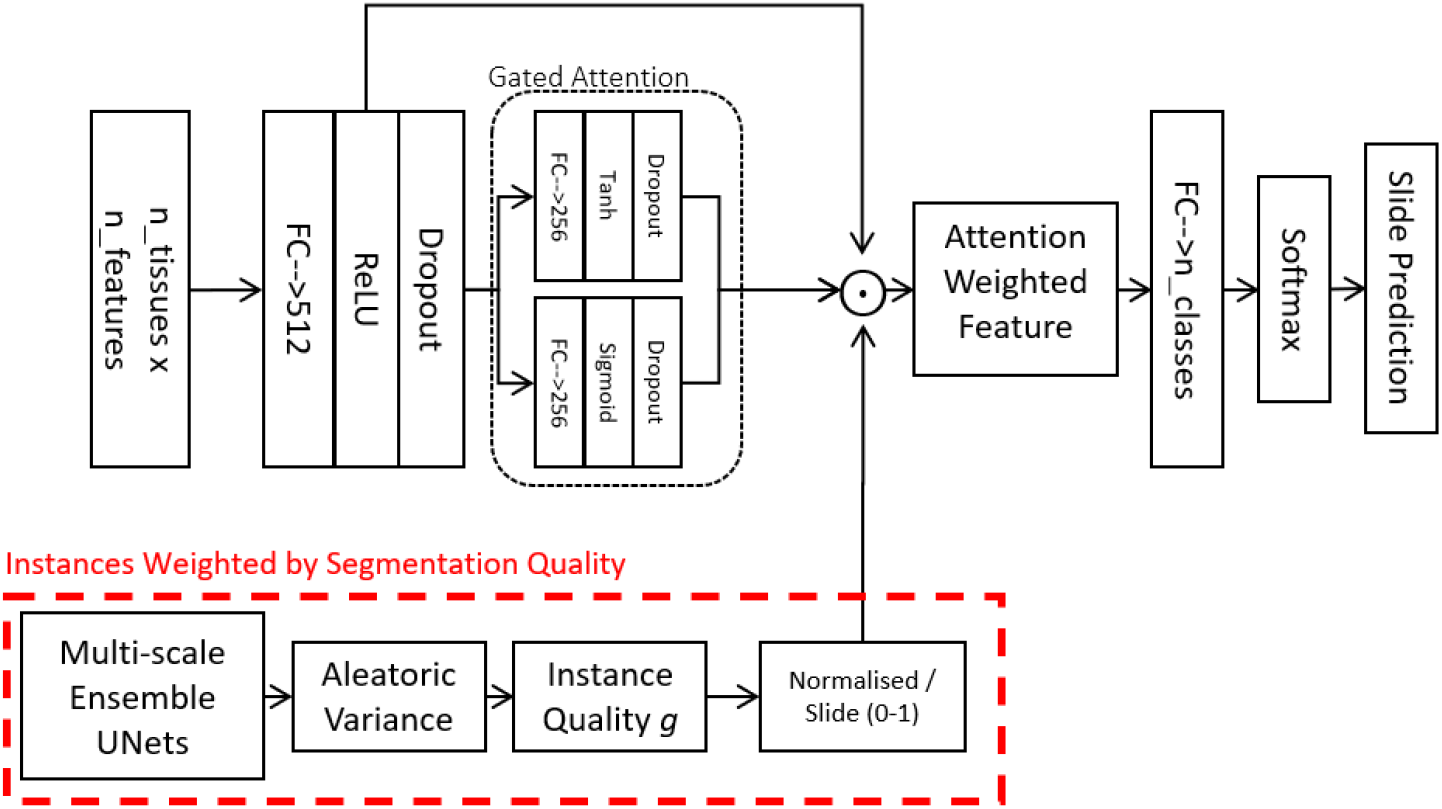
Attention Model. this model is used for combining feature vectors from a variable number of tissues into a single vector describing a slide. The gate attention module learns which instances are relevant for the bag-level prediction tasks. Optionally, in addition to the learned attention, we also weight instances by their segmentation quality and how much the instances resemble our locally-delineated training examples.

In practice, however, in many biomedical datasets, the number of instances in each bag could be greater than the number of bags, making it very difficult to train a reliable attention mechanism. The attention network may also fail to assign a meaningful score for instances that do not resemble any of those from the training examples. To address this challenge, we propose to include an additional factor **g** into the weighted average. The weight *g*_*k*_ over an instance *k* can be described as a confidence score of the neural network on an instance indicative of its resemblance to the training data. Such a score can be derived using probabilistic predictions from BNNs (Gal and Ghahramani, 2016; Blundell et al., 2015; Kendall and Gal, 2017). As we have a plethora of delineated tissues, we decided to obtain **g** using our UNet ensemble (described in Section 2.2). To account for the Bayesian uncertainty of different instances, we simply incorporate **g** into the attention as follows:

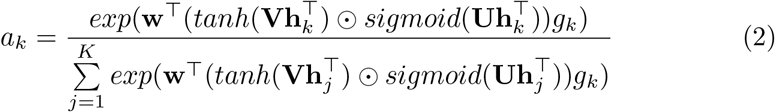

### 2.1. Datasets

Two main datasets are used in this study. A breakdown of the datasets is summarised in Table 1. The datasets contain slides stained using PAS and SR.

**Table 1:** Datasets. Slides were digitised by at least 4 different scanners. All slides from the NMP dataset, 169 QUOD-PAS slides, and 20 QUOD-SR slides were digitised using a Hammatsu scanner at 40x (0.22mpp). The other QUOD slides were digitised using a Glissando Desktop scanner at 40x (0.27mpp). The total area (*mm*^2^) selected for tissue delineation from each dataset is shown in the right-most column. (Note: 65 tiles contain only annotations of cell nuclei are not included in the table.)

| Dataset / Stain | Donors | Kidneys | Biopsies | Slides | Tiles | Delineated |
| --- | --- | --- | --- | --- | --- | --- |
| QUOD → PAS | 348 | 373 | 394 | 414 | 85 | 253 |
| QUOD → SR | 173 | 195 | 195 | 215 | 20 | 20.1 |
| NMP → HE | 35 | 35 | 169 | 169 | 240 | 400 |
| NMP → PAS | 4 | 4 | 23 | 26 | 12 | 5.89 |
| NMP → SR | 4 | 4 | 22 | 26 | 20 | 9.69 |
| Native → PAS | 12 | 12 | 12 | 12 | 26 | 235 |
| TCGA → HE | 11 | 11 | 11 | 11 | 22 | 282 |

While PAS is a routine stain for renal biopsy assessment, SR could be promising for the computational quantification of fibrotic tissues. SR is normally viewed under polarised light (Grimm et al., 2003) for maximum signal-to-noise ratio, but attempts to quantify the extent of fibrosis under unpolarised light have also been shown to be highly reproducible (Farris et al., 2011).

The QUOD Dataset consists of paraffin-embedded 22mm pre-implantation half-core needle biopsies from the *Quality in Organ Donor* Biobank^1^, a national multi-centre UK-wide bioresource of deceased donor clinical samples procured during donor management and organ procurement. These biopsies were from a larger cohort of cases where both kidneys from the donor have been transplanted and yielded similar outcomes (12-month eGFR) in both recipients. This cohort selection criteria allow us to reduce the importance of recipient-related factors amongst other variables influencing transplant outcomes. Clinical parameters of the QUOD dataset are listed in the Supplementary Materials (Table S6). We have received biopsies from n=354 donors from this dataset. Histology slides prepared from pre-implantation biopsies from n=348 donors (373 recipients) were stained in PAS and n=174 donors (174 recipients) were stained in SR. 180 donors had biopsy sections that were only stained with PAS but not SR and 6 donors were stained vice versa.

A characteristic of these QUOD biopsies is that they are very small for two reasons. Firstly, a small needle is used to minimise bleeding complications after transplant. Secondly, biopsies were halved in length as the other halves were used for other assays. The majority of slides do not contain enough tissues for full assessment according to the Banff criteria (Racusen et al., 1999) which states that ≥ 7 glomeruli and ≥ 1 artery is necessary for assessment. (Distributions of glomeruli and arteries available in Supplementary Materials Figure S3 and S4.) Slides that contain no arteries or *<* 7 glomeruli are only partially assessed. Hence only 90 PAS slides had received a full Remuzzi score. Several slides containing only fractions of an artery also received Remuzzi artery score. A large proportion of tissues also suffer from artifacts such as forceps compression, folding, or duplication of serial sections.

The NMP dataset^2^ originated from an organ normothermic perfusion experiment (Weissenbacher, 2018; Weissenbacher et al., 2019) from 35 donors. The slides were 18 gauge 33mm core needle biopsies obtained from deceased donor kidneys that were discarded as deemed unsuitable as transplants for mechanical reasons. These kidneys were placed into a normothermic machine perfusion (NMP) system. Biopsies were obtained at different time points during NMP hence most samples suffered from notable ischemic damage. Most of the slides (n=169) were stained with Haemotoxylin and Eosin (H&E) but had been computationally converted to PAS stain using a CycleGAN (Zhu et al., 2017) with image quality largely indiscernible by our collaborating pathologist. From the same dataset, we also have a smaller number of slides stained directly with PAS and SR (n=26 from 4 donors for each stain).

Apart from the two main datasets, we have also included additional slides from native biopsies ^3^ in PAS (Supplementary Section S5) and slides from The Cancer Genome Atlas (TCGA) stained in H&E. These slides were solely used for strengthening the segmentation algorithms (Section 2.2) to ensure it would generalise well to unseen data.

From each dataset, we manually marked out a number of rectangular tiles to delineate different tissue compartments - tubules, glomeruli, vessels, and cell nuclei. The number of delineated tissues for each dataset is shown in Table 2. Delineating the outline of tissues is a laborious task. To make the task scalable, initial annotations were performed by a engineer with limited training in pathology. Thus, we only have the coarse classification of these tissues. For instance, proximal tubules are what is relevant for assessment but a notable portion of objects marked as “tubules” were actually distal tubules or the collecting duct. Objects marked as “vessels” consist of a mixture of arteries, arteriole, and veins; some casts might be misidentified as sclerosed glomeruli. The boundaries of glomeruli were inconsistent regarding the inclusion of the urinary space. Owing to the small size of many biopsies, tissues that were truncated near the edge of the biopsies were also delineated as long as they were human-recognisable. A subset of these tissues was cross-checked by our pathologist (Details in the Supplementary Materials Tables S2 and S3).

**Table 2:** Overview of tissue instances delineated. This table gives an overview of the number of tissue instances delineated by hand.

| | Number of Instances | | | Area ($mm^2$) | | |
| --- | --- | --- | --- | --- | --- | --- |
|  | Tub | Glom | Vessels | Tub | Glom | Ves |
| QUOD → PAS | 2145 | 323 | 119 | 4.68 | 3.87 | 2.60 |
| QUOD → SR | 889 | 24 | 22 | 2.03 | 0.309 | 0.254 |
| NMP → HE | 13103 | 581 | 175 | 31.8 | 7.39 | 4.37 |
| NMP → PAS | 753 | 25 | 40 | 1.64 | 0.270 | 0.236 |
| NMP → SR | 1311 | 38 | 17 | 2.70 | 0.411 | 0.493 |
| Native → PAS | 1156 | 226 | 55 | 3.62 | 4.15 | 1.11 |
| TCGA → HE | 1739 | 422 | 143 | 5.84 | 7.53 | 9.49 |

The tiles selected for tissue delineation may contain a mixture of tissue-containing and blank areas and are of different sizes and aspect ratios such that it covers a diverse range of tissue morphology. As the number of tubules is far greater than the number of glomeruli and vessels, in 165 out of 425 of the tiles (967 out of 1210*mm*^2^ in terms of area) listed in Table 1 we only delineated the glomeruli and vessels but not tubules.

### 2.2. Tissue Segmentation

Segmenting tissues according to functional compartments allows us to incorporate known visual features into histology analysis and maximises the interpretability of predictions made by algorithms. Tissue segmentation was performed using an ensemble of UNets (Ronneberger et al., 2015). As we have a varying number of annotations available, we chose to use a different number of UNets for PAS and SR-stained slides.

For segmenting tissues in PAS-stained slides, we used a total of 13 models as follows: 2 models to segment cell nuclei at 0.44 microns-per-pixel (mpp); 2 models to segment tubules, glomeruli, and vessels at 0.44mpp; 3 models to segment tubules and glomeruli at 0.44mpp; 3 models to segment glomeruli and vessels at 0.88mpp; 3 models to segment glomeruli and vessels at 1.76mpp. Models that process identical tissue classes at the same magnification were trained on a different train:validation (4:1) split. These UNets were trained and validated using tissues delineated from the NMP, TCGA, and native biopsy slides.

For SR-stained tissues, we used 4 models as follows: 2 models to segment tubules, glomeruli, and vessels at 0.44mpp; 2 models to segment the same tissues at 0.88mpp. Cell nuclei are not segmented as they are not visible under SR.

To process the test data, we implemented the ensemble of UNets with dropout to simulate Bayesian Neural Networks (Gal and Ghahramani, 2016; Kendall et al., 2015) with the same hyperparemters as Tam et al. (Tam et al., 2020). The motivation for using BNNs is that they generally output predictions where the soft values are more representative of the probability of a correct prediction. However, in cases where uncertainties are data-limited (aleatoric uncertainties) rather than model-limited (epistemic uncertainties), we find that there could still be notable discrepancies between the predictions and the actual probabilities. In particular, if the relevant class is rare or looks very different in the test data, it could lead to under/over-confident predictions. Thus, we propose to correct the predictions using a data-driven approach as shown in Equation 3:

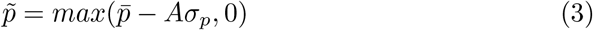

Where 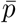 is the mean output from the neural network ensemble; *σ*_*p*_ is the standard deviations from the ensemble over a single pixel; *A* is a constant to be empirically determined from the training data; 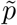 is the corrected probabilistic output from the network ensembles, which is clamped to a value above zero. As we shall see in Section 3.1, this serves to remove pixels that are overconfident and would help to suppress false-positive pixels caused by artifacts.

From the ensemble-averaged (Equation 3) segmentation maps, we obtain tissue instances using the max-flow-min-cut (Boykov and Funka-Lea, 2006) algorithm. Individual tissues are cropped from the original slide with 1.32*µm* padding on each side. In order to perform localised diagnostics based on individual tissues, areas outside of the tissues are blurred. This helps to prevent extra-tissue regions from contributing to visual features at later steps.

In our case, we have chosen to derive **g** from the UNet ensemble. For a slide with *K* tissue instances, the weight assigned to the instance *k* is given as:

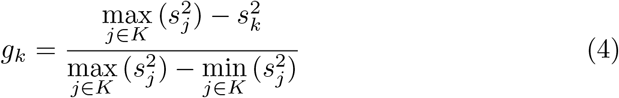

*s*_*j*_ is the mean value of *σ*_*p*_ over the segmentation mask for instance *j*. If all UNets from the ensemble predicts similar values for the pixels within instance *k, g*_*k*_ would have a value close to 1. Otherwise, high discordance between different UNets would result in *g*_*k*_ close to 0.

More details regarding the implementation of tissue segmentation is detailed in Section S3 under Supplementary Materials.

### 2.3. Extraction of Histological Features

We aim to demonstrate (i) the benefits of using handcrafted features to augment deep features and (ii) how extracting features from functional tissue structures can boost performance and interpretability in multi-instance learning settings. As there are currently very few studies that quantitatively assess how individual histological features correlate with physiologically relevant measurements, we tested a wide range of features in our study including both handcrafted and deep features. Using the aforementioned workflow, we extracted a number of histological features from our slides from tissues. We designed handcrafted features that comprise tissue morphological descriptors, colour, texture, as well as second-order features such as how colour/texture are distributed with respect to the tissue compartment. The majority of handcrafted features were designed with one tissue type in mind but implemented across all tissue classes such that the feature vector is the same length for all tissue types.

Some of these features are designed to reflect visual changes of tubules that have undergone chronic or acute injuries. For PAS-stained slides, cell nuclei are typically visible within each tissue so their colour and distribution may also shed light on the state of the biopsy. In proximal tubules, darker nuclei in epithelial cells may be a feature of mitosis and cellular repair; whereas cell nuclei located far away from the boundary of proximal tubules may signify cell dropout or cytoplasm expansion which is a feature of acute tubular injury (Pieters et al., 2019; Solez et al., 1993). As the number of nuclei in each tissue is variable, the values are pooled together at every tenth percentile. This has an advantage over max-pooling of being less sensitive to artifacts/falsely detected cell nuclei.

The full list of handcrafted features used in this study is shown in Table S7. Several features are derived from the distribution and colour of segmented cell nuclei. However, we recognise that some tissue compartments may not have any nuclei so these will lead to missing feature values. To prepare our data for machine processing, missing values are imputed with the mean value from the rest of the datasets. Some slide-level information, such as the total area of the biopsy, is appended to the feature vector of each individual tissue. In total, this resulted in 98 unique handcrafted features for the PAS-stained slides and 40 features for the SR slides.

Figures 3 and 4 show selection of tissue examples with close to minimum / maximum feature values from the QUOD/PAS slides. In addition to handcrafted features, we also experimented with features from several established deep neural networks. Deep features are obtained from the same patch (with surroundings blurred) as the handcrafted features at 0.44mpp. The crops were not resized before we fed them into neural networks as we want objects of the same physical size to elicit the same filter responses. Fullyconnected layers of neural networks were replaced by adaptive average pooling, resulting in a single 1D feature vector for each tissue. Each feature is normalised to unit variance with zero mean over our datasets to speed up convergence during training.

**Figure 3:**
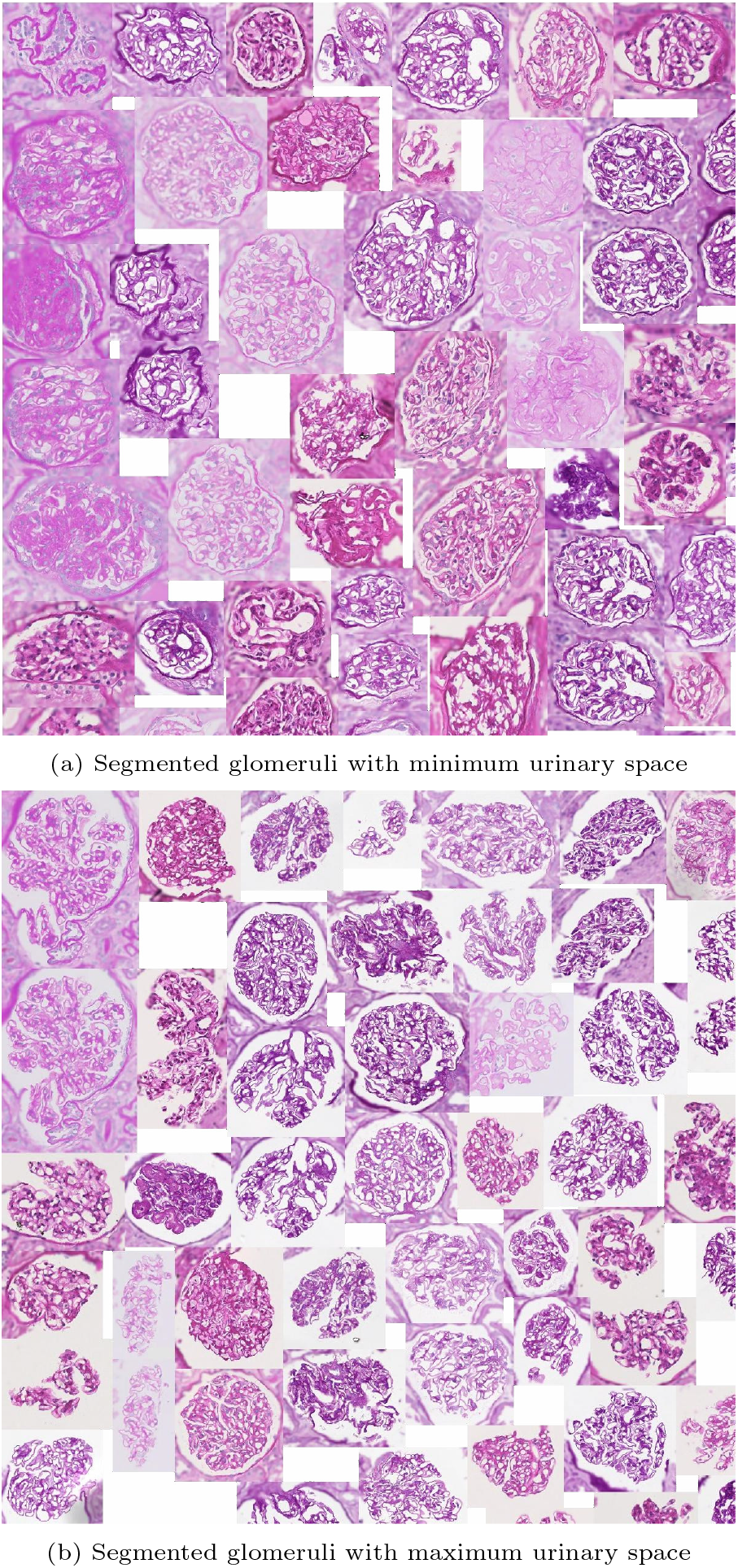
Glomeruli with (a) minimum / (b) maximum urinary space area. Examples are algorithmically selected from the entire QUOD dataset. Visual differences of individual handcrafted features can be easily interpreted by inspecting the collection of tissues with low vs high values. Note that regions outside the tissue are blurred

**Figure 4:**
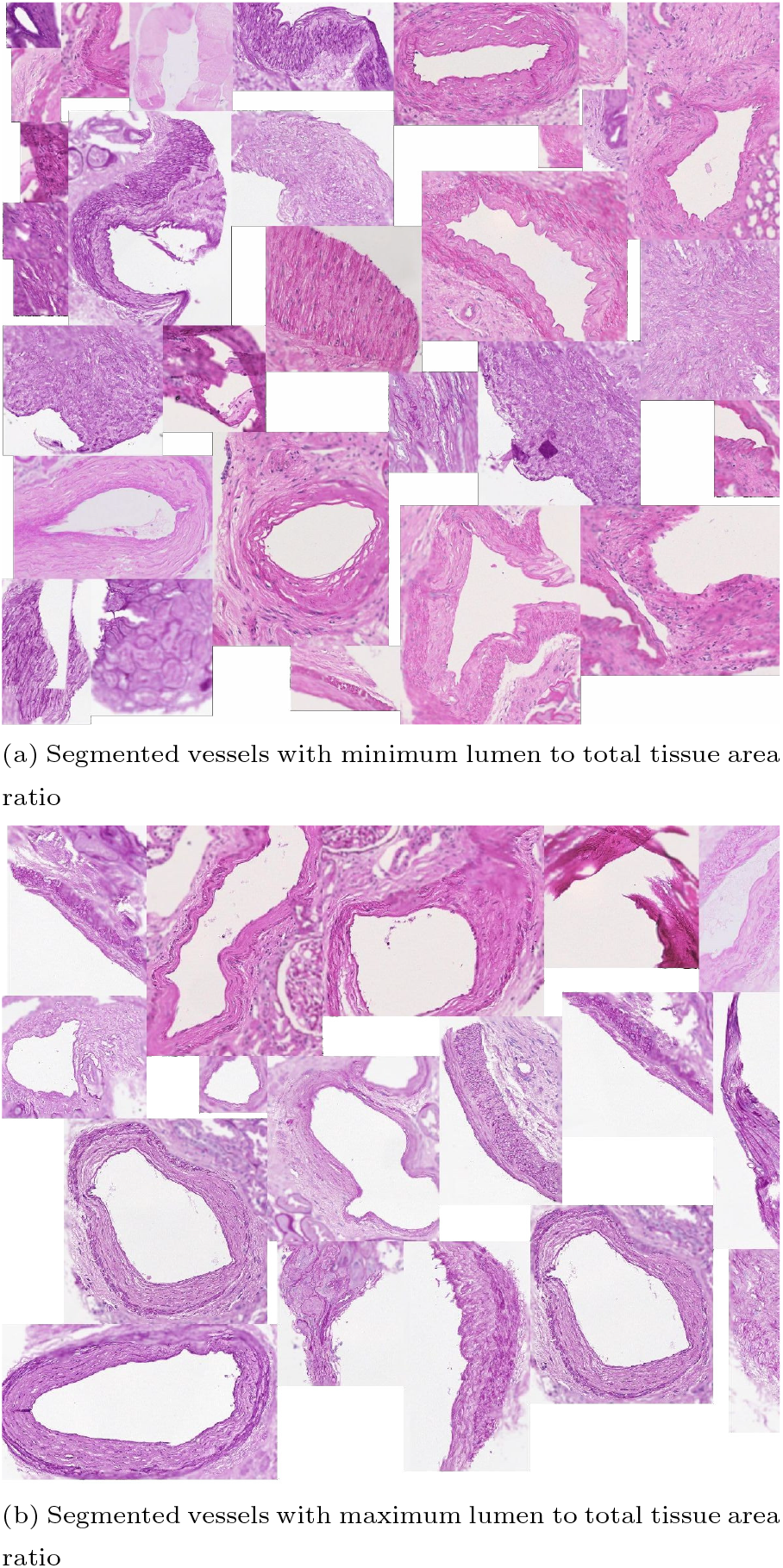
Vessels with (a) minimum / (b) maximum ratio between the lumen area to total vessel area. This exemplary feature corresponds to one of the Remuzzi Score criteria. Examples are algorithmically selected from the entire QUOD dataset. We choose to calculate the ratio of areas instead of diameters to avoid geometric template fitting as most vessels sections are not round.

### 2.4. Predicting Slide Level Labels

Several different physiologically relevant measurements are available for our datasets.

A subset of PAS-stained slides has been assessed by an experienced pathologist (blinded to the donor characteristics and outcome) to determine the extent of histological changes. Slides are graded according to the Remuzzi criteria (Remuzzi et al., 2006) based on the severity of Tubule Atrophy (Remuzzi TA), arterial and arteriolar narrowing (Remuzzi A), glomerular global sclerosis (Remuzzi G), and interstitial fibrosis (Remuzzi IF). In addition, as part of the assessment routine, biopsies are also graded for Acute Tubular Injury (ATI). The distribution of the assessed grades can be found in the Supplementary Materials (Figure S1).

For labels with insufficient cases for training or testing, we regrouped the most severe cases until there is enough donors for cross-validation to reduce class imbalance. As a result, Remuzzi G, TA, and IF become a binary classification task; whereas labels for ATI are regrouped to either two or three (0-2) grades instead of four grades from the original assessment.

Apart from eGFR, the QUOD dataset also contains binary labels regarding whether the recipient has suffered from DGF. DGF may have origins from a variety of diagnoses. While ATI is one of the known leading culprits, there are also other causes such as T cell-mediated rejection, antibody-mediated rejection, and acute calcineurin toxicity (Kers et al., 2018; Rolak et al., 2021). While it may not be possible to detect some recipient-related factors (such as rejection) or surgical causes (such as anastomosis) from histology, there may be subtle or localised acute lesions within small regions of some biopsies. A method based on multi-instance learning may have a chance of detecting these localised changes missed by human inspectors or not meeting histological thresholds of established grading criteria.

Combinations of handcrafted histological features and deep features are used as input for our multi-instance soft attention model (Figure 2). The length of training is determined using the validation AUC at 40 epochs after the metric becomes stagnant. A weighted sampling approach was used to increase the frequency of sampling the rarer classes.

To reduce bias, hyperparameters of our attention model are tuned based on a trivial task. Slides are labelled by whether they contain enough glomeruli for assessment. Slides with *<* 7, 7 − 9, and ≥ 10 unique glomeruli are separated into three different classes. Hyperparameters are searched using HyperOpt (Bergstra et al., 2013) and remain fixed for other tasks in order to be able to compare different featuresets fairly. Examples of hyperparameters explored include the choice of gated/un-gated attention network, size of network layers, and regularisation weight.

Because the QUOD slides were obtained from real transplant settings, there were several challenges to developing an algorithmic workflow based on these slides. Firstly, these slides consist mostly of kidneys with little chronic damage - the dataset is biased due to pre-transplant donor screening and the distribution of biopsies with pathological changes is highly imbalanced. Secondly, the majority of biopsies are inadequately small that they do not meet the Banff criteria (Racusen et al., 1999). For predicting assessment grades given by pathologists, we only included slides with enough tissues for each grade. As for the prediction of DGF, we found that it was necessary to include slides of all sizes in order to achieve at least one donor per label per cross-validation. Cross-validation splits are subjected to the constraint where all slides from each donor remain in the same training/validation/test set.

## 3. Results and Discussion

Experimental results are presented in four sections. Section 3.1 briefly describes the segmentation results on the QUOD dataset. In Section 3.2, features are extracted based on segmented tissue instances and are used to predict slide labels. We also compare classification performance between different featuresets across several tasks. Section 3.3 shows a pilot visualisation scheme of the proposed workflow. Finally, Section 3.4 presents results showing segmentation uncertainty can be used to improve prediction performance.

### 3.1. Segmentation Results

As per earlier experiments, we found that UNets that were trained on the same magnification tend to produce false-positive segments in the same areas of the test data regardless of how dataset splits, weight parameters, input tile size, and regularisation are initialised. These false positives were likely caused by tissues with morphology not present in the training data. Combining multiple magnifications according to Equation 3 has helped to remove most of these false positives. Figure 5 shows the segmentation examples from individual models and the combined soft prediction. It can be seen that most artifacts from individual UNets (b-e) are no longer visible once combined (f). Although it may be sufficient to use fewer UNets in the ensemble to reduce the amount of computational work, for the purpose of this experiment we are interested in the rest of the workflow where segmentation is not a limiting factor. More details on tissue segmentation is described in Section S3.

**Figure 5:**
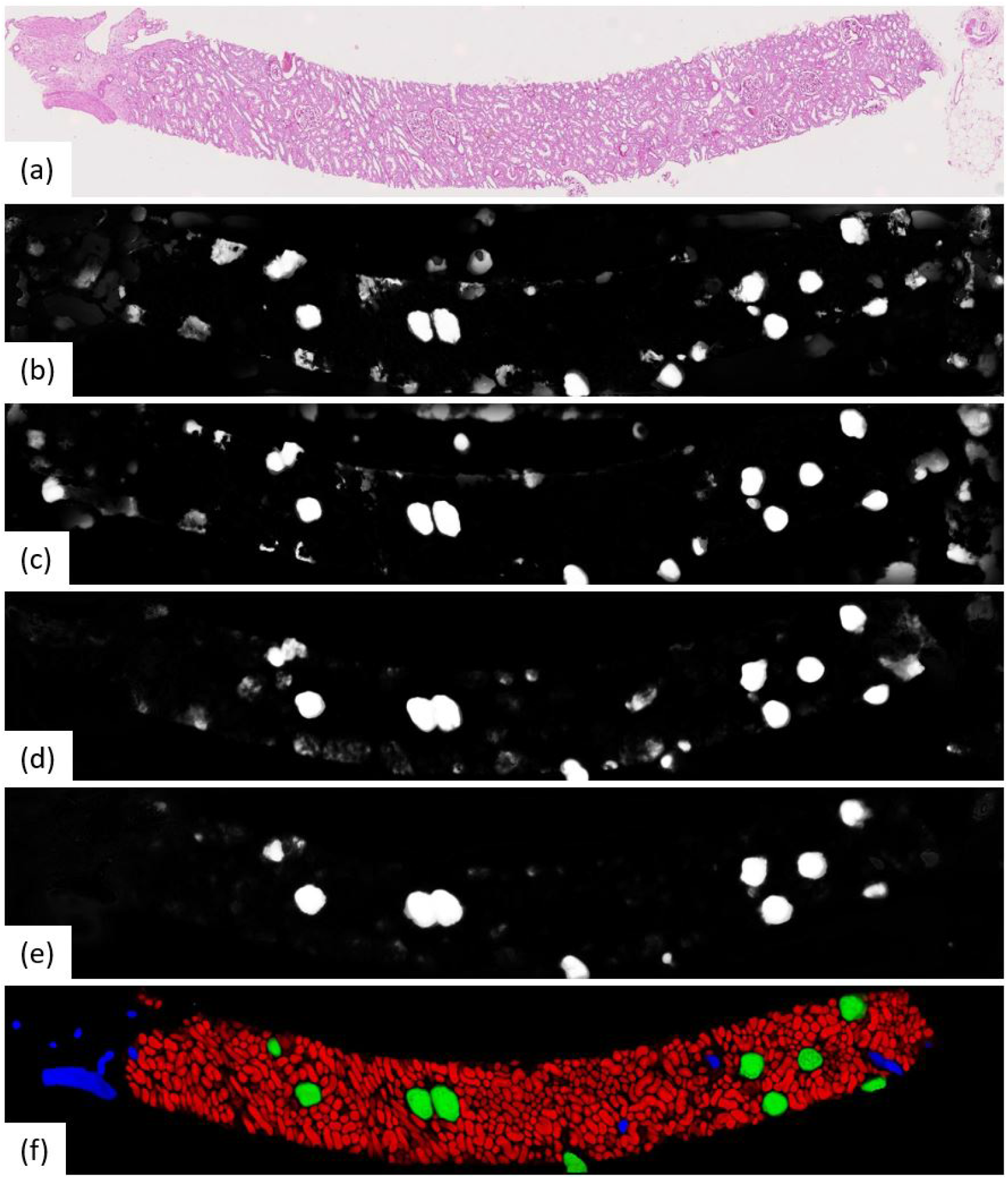
Output from the UNet ensemble. (a): Original PAS-stained slide; (b)-(e): Softmax predictions of glomeruli from single UNets at different magnifications segmenting specific tissue classes. (b) 0.44mpp, all tissue classes; (c) 0.44mpp, tubules + glomeruli; (d) 0.88mpp, glomeruli + vessels; (e) 1.76mpp, glomeruli+vessels; (f) Ensemble-combined predictions showing tubules, glomeruli, and vessels in red, green, and blue respectively.

### 3.2. Performance of Different Featuresets

In this section, the soft attention models are implemented directly without accounting for the segmentation quality (Figure 2) of the tissues.

We compared predictive performance using a variety of featuresets as the input to the soft attention models. In order to avoid the need to set up an arbitrary threshold, we reported results in the form of ROC and PR curves. Curves are weighted by the number of tissues/patches in each slide to reduce the noisy impact from biopsies that do not meet the Banff criteria.

To draw a conclusion of the optimal methodology, we have calculated not just AUCs but also their variability. Reported ROC-AUC values of the soft attention model in all tables in this paper averaged across 5 fold cross-validation test set (3:1:1 training/validation/testing) and 5 different seeded weight initialisation (total 25 models). Multi-class models are macro-class-averaged. We reported the unbiased standard error of the mean AUC to ensure that comparisons are meaningful and to account for data and model noise. Standard errors are calculated as if the different prediction tasks are independent. In addition, because training was performed on mixed datasets, neural networks may have learned to look for “shortcuts” (eg. classification based on staining protocol rather than pathological changes) instead of performing the main task. Thus, all AUC values are evaluated based on only the QUOD slides.

Deep features are extracted from a number of neural networks, including several networks pre-trained with ImageNet (ResNet50 (He et al., 2016), VGG16 (Simonyan and Zisserman, 2014), InceptionV3 (Szegedy et al., 2016)), two networks trained using cropped image patches (ResNet50 and a Variational AutoEncoder (Kingma and Welling, 2013)), and ScatterNet (Bruna and Mallat, 2013). Details of these features is summarised in Table 3 with further description in the Supplementary Materials Table S8. Features extracted from the segmented tissues are given the prefix “Tissue”. We have also extracted features using fixed-sized rectangular tiles - these are given the prefix “Tiles” in the table. While handcrafted features can be calculated for individual tissues, there is no intuitive way to do so for rectangular tiles as they may contain a varying number of tissues. A breakdown of the predictive performance of some of these featuresets for different tasks is shown in Table 4.

**Table 3:**
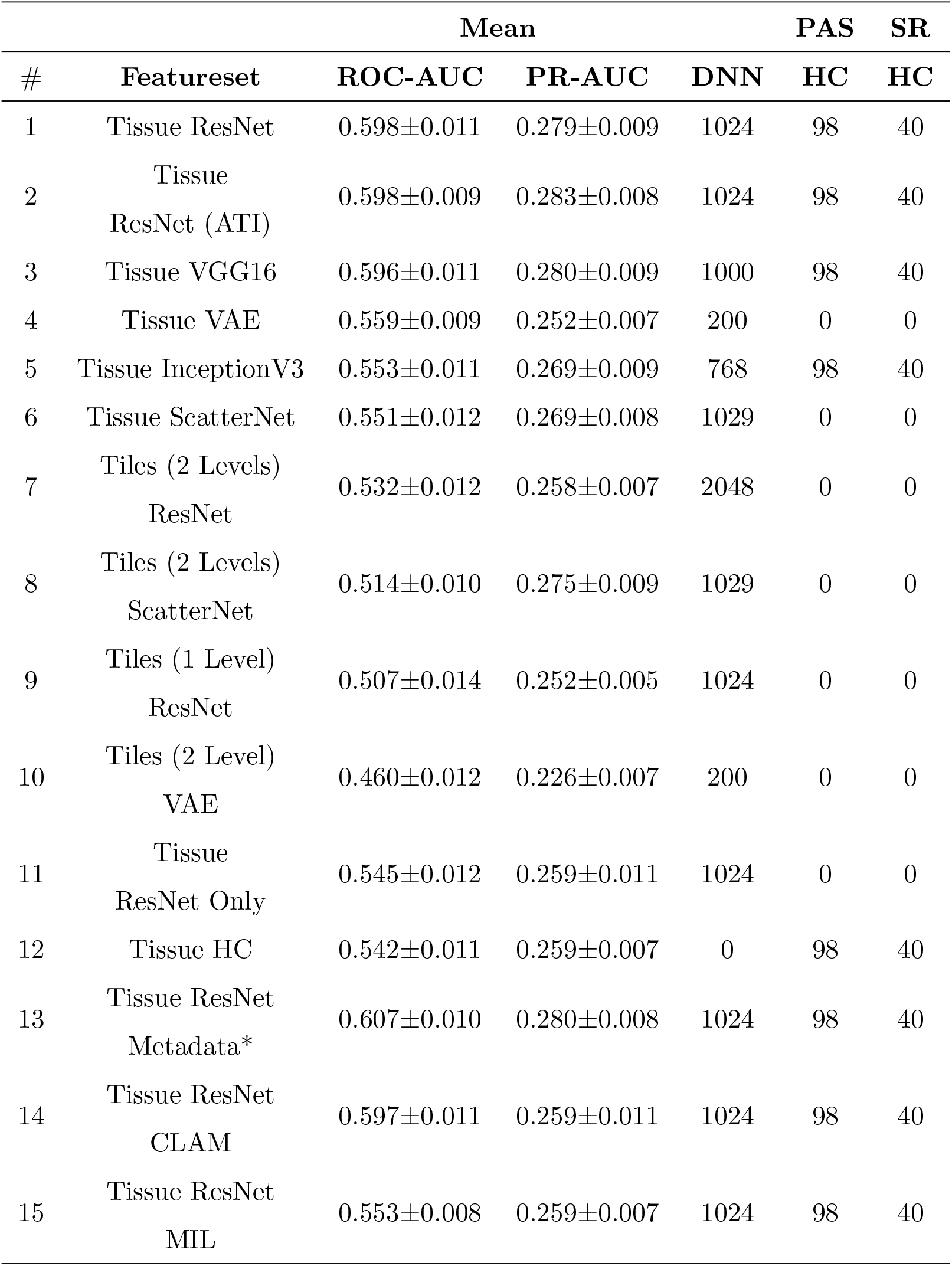
Overview of featuresets. Featuresets are made by concatenating deep neural network (DNN) and handcrafted (HC) features. We have also tested adding donor/recipient metadata (*Supplementary Materials Section S4) as feature vectors as shown in row 13. Mean AUC values over multiple tasks are shown. Detail breakdown of AUC values of some of these featuresets is shown in Table 4. Qualitative description of these featureset is also available in Table S8.

**Table 4:**
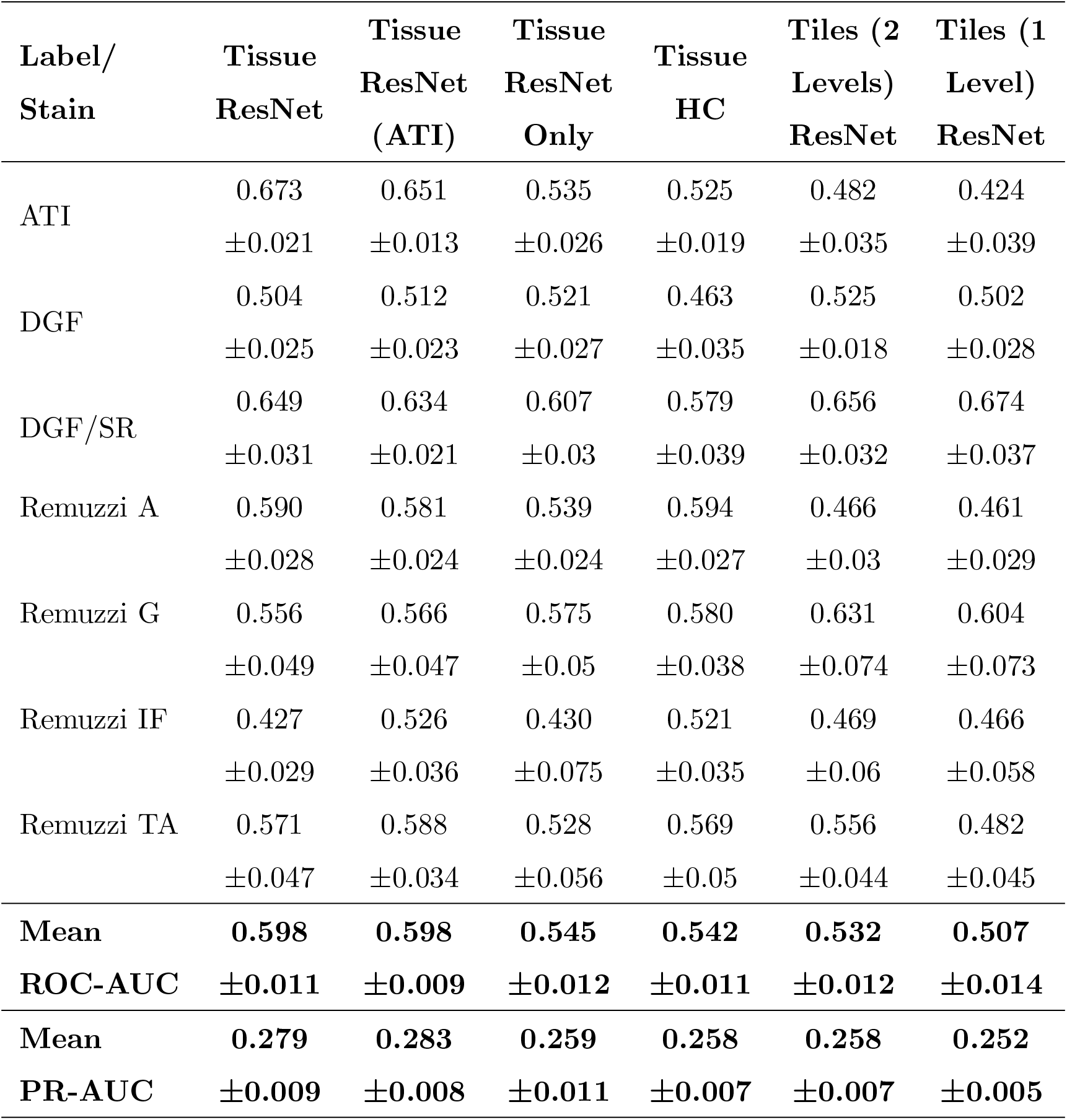
Overview of AUC values of predictions based on different featuresets (columns) and prediction tasks (rows). The second last row shows the inverse-variance weighted mean of the various tasks above. Columns are arranged in descending order of the mean ROC-AUC. It can clearly be seen that features extracted from tissues (column prefix “Tissue”) perform better than features extracted from fixed-sized rectangular tiles (column prefix “Tiles”). Note also that combining deep features with handcrafted features (Tissue ResNet, *AUC* = 0.598) resulted in better performance than using only either deep features (Tissue ResNet Only, *AUC* = 0.545) or handcrafted features (Tissue HC, *AUC* = 0.542).

| **Label/**<br>**Stain** | **Tissue**<br>**ResNet** | **Tissue**<br>**ResNet**<br>**(ATI)** | **Tissue**<br>**ResNet**<br>**Only** | **Tissue**<br>**HC** | **Tiles (2**<br>**Levels)**<br>**ResNet** | **Tiles (1**<br>**Level)**<br>**ResNet** |
| --- | --- | --- | --- | --- | --- | --- |
| ATI | 0.673 | 0.651 | 0.535 | 0.525 | 0.482 | 0.424 |
| | $\pm 0.021$ | $\pm 0.013$ | $\pm 0.026$ | $\pm 0.019$ | $\pm 0.035$ | $\pm 0.039$ |
| DGF | 0.504 | 0.512 | 0.521 | 0.463 | 0.525 | 0.502 |
| | $\pm 0.025$ | $\pm 0.023$ | $\pm 0.027$ | $\pm 0.035$ | $\pm 0.018$ | $\pm 0.028$ |
| DGF/SR | 0.649 | 0.634 | 0.607 | 0.579 | 0.656 | 0.674 |
| | $\pm 0.031$ | $\pm 0.021$ | $\pm 0.03$ | $\pm 0.039$ | $\pm 0.032$ | $\pm 0.037$ |
| Remuzzi A | 0.590 | 0.581 | 0.539 | 0.594 | 0.466 | 0.461 |
| | $\pm 0.028$ | $\pm 0.024$ | $\pm 0.024$ | $\pm 0.027$ | $\pm 0.03$ | $\pm 0.029$ |
| Remuzzi G | 0.556 | 0.566 | 0.575 | 0.580 | 0.631 | 0.604 |
| | $\pm 0.049$ | $\pm 0.047$ | $\pm 0.05$ | $\pm 0.038$ | $\pm 0.074$ | $\pm 0.073$ |
| Remuzzi IF | 0.427 | 0.526 | 0.430 | 0.521 | 0.469 | 0.466 |
| | $\pm 0.029$ | $\pm 0.036$ | $\pm 0.075$ | $\pm 0.035$ | $\pm 0.06$ | $\pm 0.058$ |
| Remuzzi TA | 0.571 | 0.588 | 0.528 | 0.569 | 0.556 | 0.482 |
| | $\pm 0.047$ | $\pm 0.034$ | $\pm 0.056$ | $\pm 0.05$ | $\pm 0.044$ | $\pm 0.045$ |
| **Mean** | **0.598** | **0.598** | **0.545** | **0.542** | **0.532** | **0.507** |
| **ROC-AUC** | **$\pm 0.011$** | **$\pm 0.009$** | **$\pm 0.012$** | **$\pm 0.011$** | **$\pm 0.012$** | **$\pm 0.014$** |
| **Mean** | **0.279** | **0.283** | **0.259** | **0.258** | **0.258** | **0.252** |
| **PR-AUC** | **$\pm 0.009$** | **$\pm 0.008$** | **$\pm 0.011$** | **$\pm 0.007$** | **$\pm 0.007$** | **$\pm 0.005$** |

To compare the general utility of the different methodologies we averaged the AUCs from different tasks. ROC-AUCs are averaged with inverse variance weighting so tasks with higher prediction consistencies are weighted more. Precision-Recall (PR) curve AUCs are also given in results (Table 3) as a complementary metric to ROC-AUCs. PRs could be insightful for tasks with highly imbalanced labels. However, variances of PR-AUCs tend to be small for the more challenging tasks, so we only reported the arithmetic mean rather than the inverse variance weighted mean in the table.

The mean ROC-AUC values in Table 3 demonstrate the progress towards improving performance by using features extracted from tissue compartments versus the simplistic model that uses only features from rectangular tiles. For example, when comparing the ResNet50 featuresets (rows 1 and 7) we get *AUC* = 0.598 ± 0.011 and 0.532 ± 0.012 for features extracted from tissues and rectangular tiles respectively. These results show that our proposed approach is superior in most cases. If we look at the results at a more granular level, we will find that there are some tasks where tile features may have the potential to outperform tissue features. From the prediction task breakdown for these two featuresets (corresponding columns in Table 4), we see that tiles perform better for Remuzzi G (0.556 ± 0.049 vs 0.631 ± 0.074), DGF/PAS (0.504 ± 0.025 vs 0.525 ± 0.018), and DGF/SR (0.649 ± 0.031 vs 0.656 ± 0.032). However, the differences for these individual tasks have not yet reached statistical significance so this could be down to noise in the data.

A second observation to note is that performance is better when deep and handcrafted features are combined compared to using only deep features or only handcrafted features (*AUC* = 0.598 ± 0.011, 0.545 ± 0.012, and 0.542 ± 0.011 in respective order as seen from rows 1, 11, and 12 in Table 3). While the overall AUC values for “Tissue ResNet50 Only” and “Tissue HC” are not significantly different, from the task breakdown in Table 4 we can see that AUCs are more variable for the predictions based on handcrafted features. This could be because handcrafted features are specialised in specific tasks. For example, in our dataset, most slides that have been graded for Remuzzi A had only the minimum number of one artery, many of which are partially truncated. So it may be hard for our attention model to learn to predict Remuzzi A grades based on only deep features. Handcrafted features can help to supply complementary information based on domain knowledge. These results suggest that both types of features may have their respective advantages. Thus, implementing a hybrid approach in the workflow may be optimal for general tasks.

Furthermore, we have also attempted to compare our soft attention model with other multi-instance methods such as CLAM (Lu et al., 2021) and MIL (Campanella et al., 2019; Maron and Lozano-Pérez, 1998) as shown in rows 14 and 15 in Table 3. While soft attention has consistently outperformed MIL, comparison is more challenging for CLAM due to the extra hyperparameters involved. An extensive search over several hyperparameters using HyperOpt’s Bayesian optimisation algorithm (Bergstra et al., 2013) with the Ray Tune platform (Liaw et al., 2018) has shown that the extra clustering step in CLAM has not led to any benefits to our prediction tasks.

In additional to neural networks pre-trained with ImageNet, we have modified a ResNet50 architecture to predict ATI scores (0-3) based on localised tissue patches (Image patches with ATI distribution shown in Figure S7). This modified ResNet was trained and validated on 731 images of proximal tubules with a 4:1 split. The purpose of this model is to test whether convolution filters would better capture histopathological changes if it has prior exposure to such images. The results (*AUC* = 0.598 ± 0.011, 0.598 ± 0.009 from rows 1 and 2 in Table 3) show there is no significant difference in performance regarding how the networks were trained, suggesting the convolution filters learned from ImageNet may already be adequate for capturing diversity in renal histology.

### 3.3. Attention Visualisation

Multi-instance learning on whole slide images is not commonly trained end- to-end due to their large size. Features are usually saved onto the disk before being processed by the MIL model. Gradients from neural network feature extractors could take up an enormous amount of space, so they are discarded during the feature extraction process in real-life implementations. As a result, the most straightforward parameters that could be directly visualised are the attention values attributed to the different instances. However, visualisation of attention parameters could often be ambiguous. In the case where rectangular tiles are chosen, the resolution of the attention map is limited by the size of the tile. Localisation of the diagnostic would be poor if the tile size is too large. If the tile size is too small, there would be limited receptive field and we may risk truncating meaningful tissue structures; a hybrid approach that uses tiles at multiple scales may be complicated to visualise as interpolations may be required to fuse them together.

On the other hand, soft attention mechanism based on individual tissues allows improved diagnostic interpretability strictly confined to the anatomical boundaries of these tissues. Figure 6 show an example of how our visualisation scheme (left) compares to the standard approach (right) which uses deep features from rectangular tiles. The models producing these exemplary overlay images were both trained to predict Remuzzi G grade. Both overlays show that the networks have learned to attend to the Glomeruli. In the standard approach, we see that the tiles around most glomeruli are highlighted in red, but in many cases, it may not be clear to an inexperienced observer as to which tissues within the tiles triggered the prediction. Conversely, our approach shows clear pinpointing of the tissues of interest as the glomeruli are mostly highlighted in red whereas irrelevant tissues, including those adjacent to the glomeruli, are shaded in blue.

**Figure 6:**
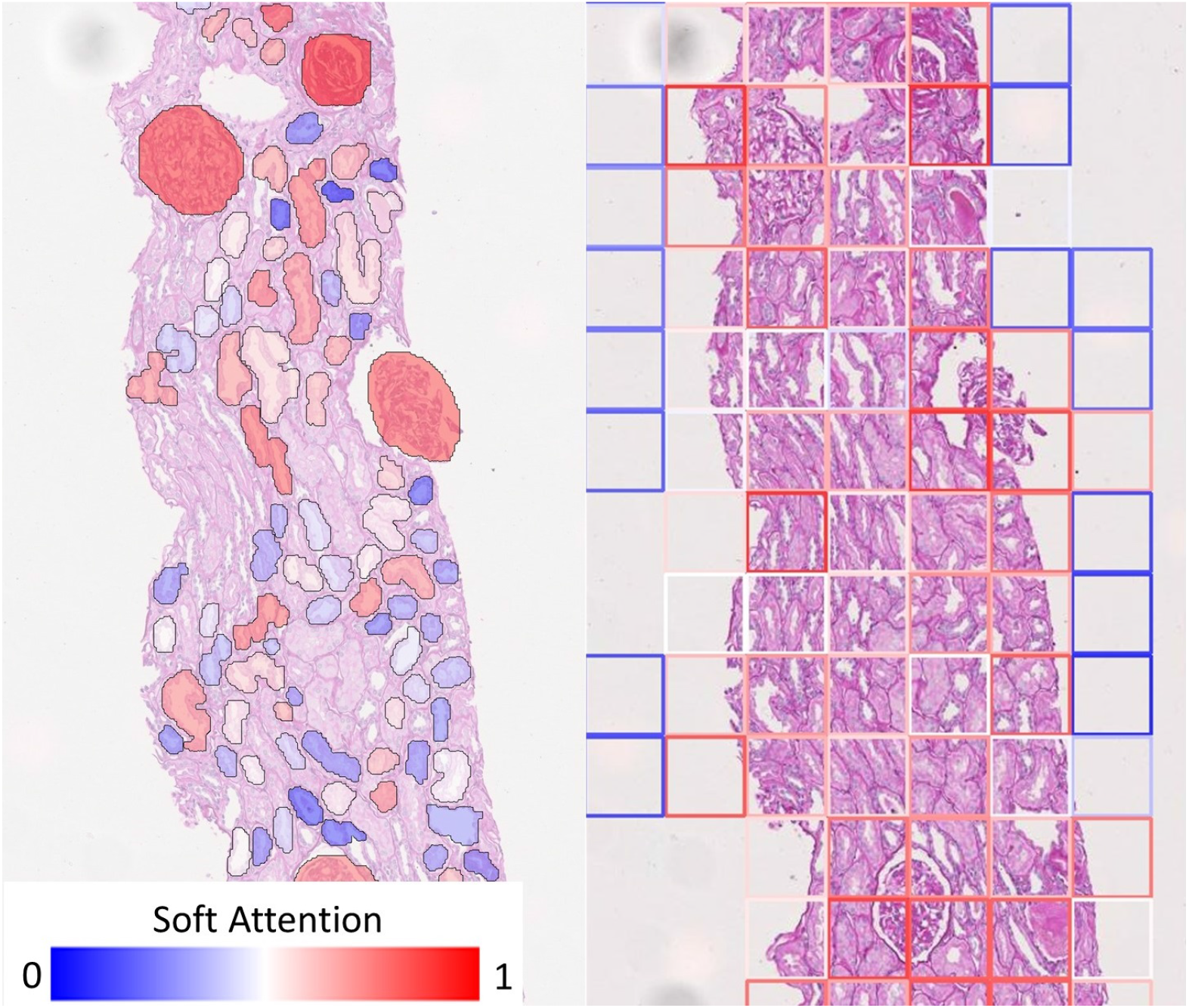
Comparison of visualisation of attention map. (Left) Our visualisation scheme highlights individual tissue instances relevant to the prediction. (Right) If rectangular tiles are used diagnostic could be ambiguous as the tile boundaries do not generally convey any diagnostic meaning. This makes it hard to pinpoint the offending tissue if they are much larger or smaller than the tile. While it may be possible to produce a smoother heatmap by using overlapping tiles, this will merely be a visual gimmick - the resolution of the attention map cannot be improved because information would have already been lost.

Saliency maps within individual instances can be produced if greater clarity is desired at the cost of processing time. In implementations where only deep features are used, it may be possible to produce saliency maps directly by chaining up feature extractors with the soft attention model while keeping track of gradients in only a small number of instances. If handcrafted features are included, however, direct localisation to the image space would not be possible using the original network. In these cases, heatmaps can be produced with an ad-hoc network trained using only the relevant tissues (as given by the soft attention model) to predict slide-level labels.

We demonstrate the overlay of saliency map based on a soft attention model trained on ResNet50+handcrafted features to predict binary ATI grades as this is a case with high AUC (corresponding model in Figure 9). Using the attention values and slide labels, we trained a ResNet18 model to predict ATI slide labels using individual tissues as inputs. The network is trained using L2 loss where different instances are scaled by outputs from an attention model which learns the same task. All instances are used for training this ResNet18 model but instances with low attention scores contribute to smaller loss. Figure 7 shows the result of this attempt - saliency map within each tissue is produced using an occlusion-based approach (Zeiler and Fergus, 2014) using Python package Captum (Kokhlikyan et al., 2020); whereas the outline of the tissues is colourcoded by the instance-level attention. We can see that the attention model has learnt to focus on the proximal tubules in the cortical regions of the biopsy (outlined in red). Within these tissues, there is some evidence that shows the saliency maps are highlighting areas close the boundaries where the epithelial cells are located. Apart from the occlusion-based approach, we also attempted to use Integrated Gradients (Sundararajan et al., 2017) and Noise Tunnel (Adebayo et al., 2018), but visualisation was found to be less intuitive as the saliency maps generated are too rough for the magnification we worked on.

**Figure 7:**
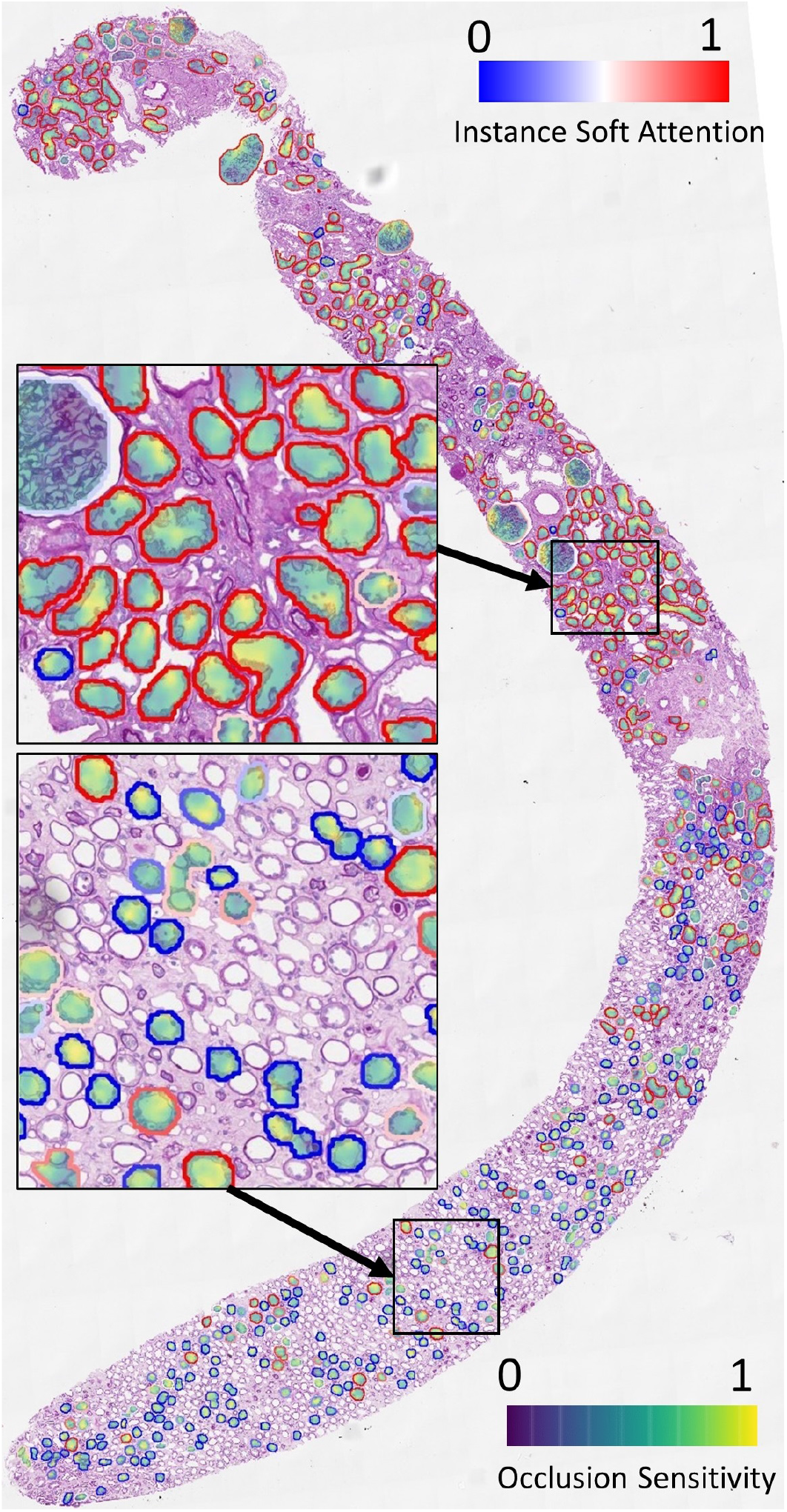
Visualisation of instance-level attention combined with saliency maps from individual tissues. Tissue boundaries are coloured by the learned instance-level soft attention value, indicating its relevance to the slide-level prediction. In this example, the slide-level label is ATI. The image clearly shows our network is able to attend to proximal tubules (mostly outlined in red). The saliency maps are produced from an occlusion-based approach using an additional network trained on individual tissues to predict the slide label.

### 3.4. Confidence-Weighted Predictions

In the previous sections, we presented results whereby tissue features were combined using soft attention models without accounting for the quality of the segmentation. We argue that the soft attention vectors learned by the models may not be always meaningful. This would happen if the instances do not resemble those in the training data, in which case the presence of the instance would only contribute to noise. We propose a scheme to take into account the quality of the instances in the form of Equation 2 and 4. Under the proposed scheme, **g** is added as a feature to the original featureset and each instance *k* is weighted by *g*_*k*_ during both training and testing time.

Figures 8, 9, and 10 show examples of ROC/PR curves on three different tasks, all of which were weighted by segmentation quality in the models. The solid plot lines show the median values and the shaded regions show the range of five bootstraps. Figure 8 is a plot of the “Trivial” task where models are trained using only ResNet50 features to classify whether a slide contains an adequate number of glomeruli. This task differs from counting glomeruli as there are duplicated tissue sections in 268 out of 612 slides (some slides contain multiple cut-throughs of the same biopsy). A naive model that only counts glomeruli would overestimate the number. We tuned our model based on this task as all slides are labelled so the dataset-induced noise would be minimal.

**Figure 8:**
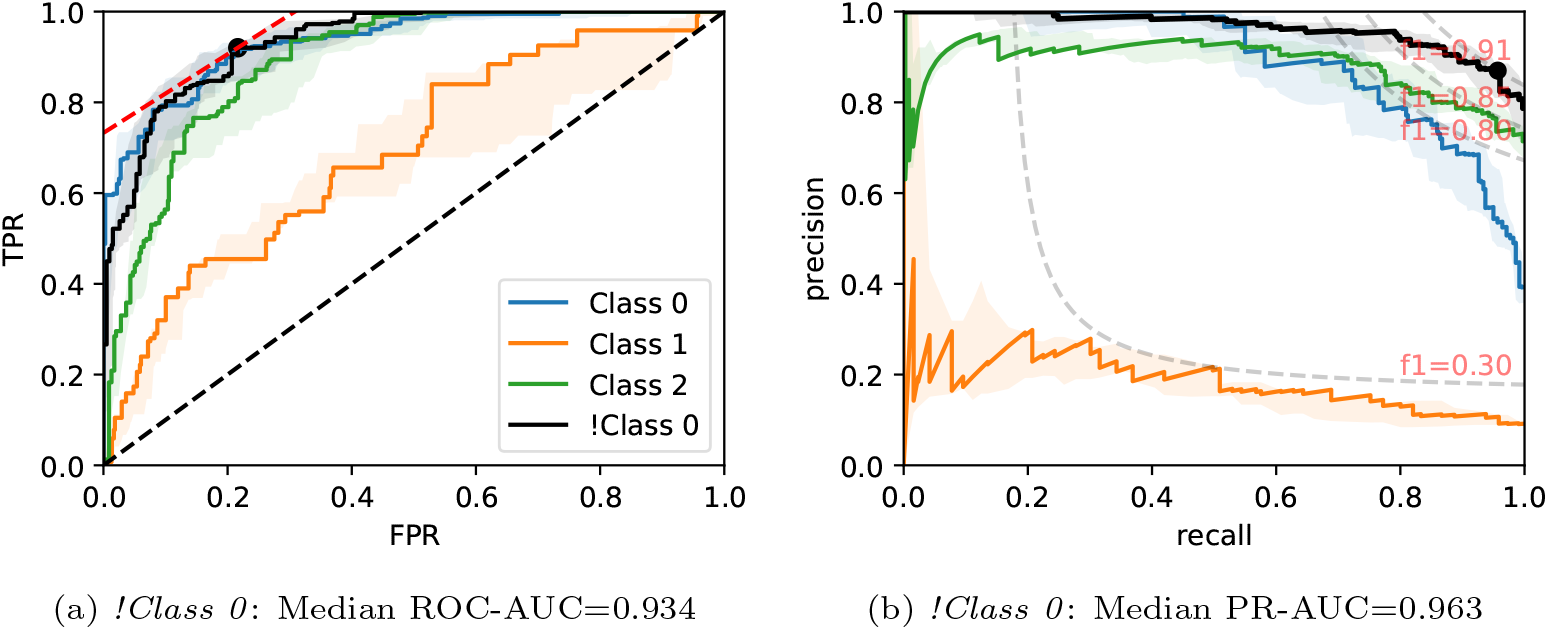
ROC/PR plots from tissue quality-weighted models predicting whether a PAS-stained slide has an adequate number of unique glomeruli for assessment (3 classes). *Class 0* : *<* 7; *Class 1* : 7 *−* 9; *Class 2* : *≥* 10; *!Class 0* : *≥* 7 glomeruli. The curves with the median AUC from 5 bootstraps are shown as solid lines. There are duplicated tissue sections in approximately 268 of the slides so models are likely to overestimate the number of unique glomeruli. The marker on the plots shows the optimal operating threshold for the respective class. ROC/PR curves for *!Class 0* represent the performance when *Class 1-2* are grouped together as a single class.

**Figure 9:**
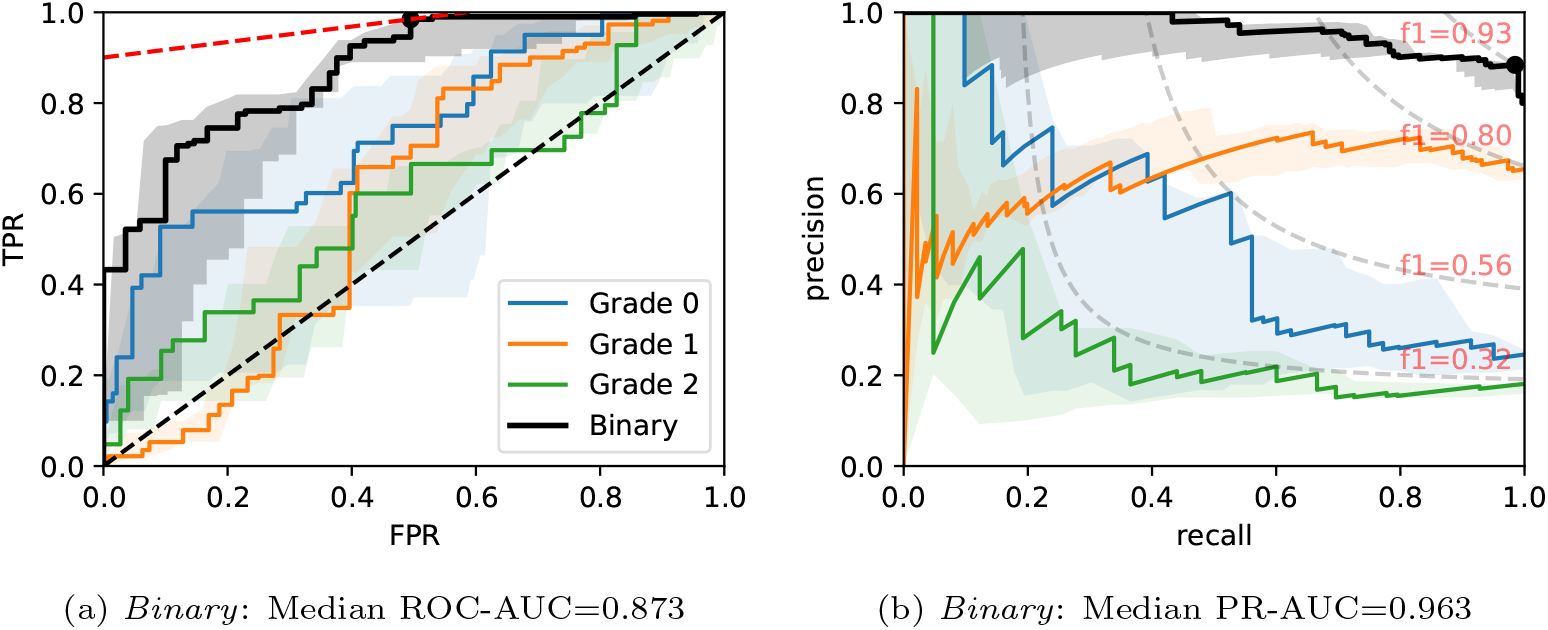
ROC/PR plots from tissue quality-weighted models predicting ATI grades based on PAS slides. Models are trained using ResNet50 and handcrafted features. Curves with the median AUC from five bootstraps are shown as solid lines. (*Grade 0-2*): Models trained to predict ATI grades 0, 1, and *>* 2; *Binary:* ROC/PR curves from models specifically trained to predict only two grades: (0, *>* 0).

**Figure 10:**
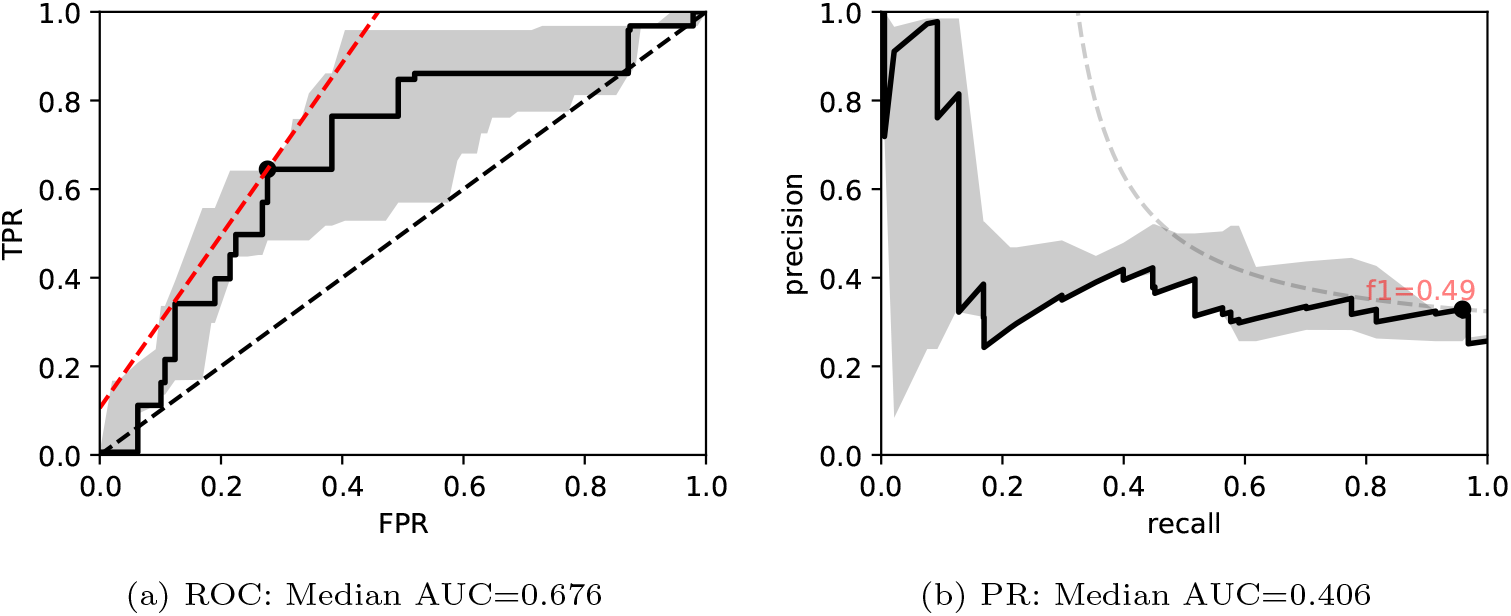
ROC/PR plots from tissue quality-weighted models predicting the presence of DGF after transplant based on SR slides. 37 out of 143 cases exhibited DGF. Models are trained using ResNet50 and handcrafted features. Curves with the median AUC from 5 bootstraps are shown as solid lines.

Figure 9 show ROC/PR curves for ATI grades where each solid line (Grade 0-2) represents the trade-offs from one-vs-other classification with models trained using ResNet50 combined with handcrafted features. If the grading is instead re-formulated as a binary task (labelled “Binary” in the plot), grouping all *>* 0 grades together and re-training the models, we get a mean *AUC* = 0.874±0.016. The optimal operating threshold can be found using the curve’s intercept with the maximum iso-accuracy line. At this threshold, we report a performance of *tpr* = 0.985 and *fpr* = 0.495.

As for DGF, we find that performances are generally higher in SR slides. Figure 10 shows the performance of the models trained on ResNet50 and handcrafted features. Based on the optimal threshold from the ROC curve, we get *tpr* = 0.645 and *fpr* = 0.277.

Table 5 shows the mean AUCs for different tasks with the proposed addition where instances are weighted by segmentation quality. Entries in this table correspond to those in Table 4. Again, comparison of individual predictive tasks is not always possible due to the limited size of the labelled data, but an improvement can clearly be seen when we average over different tasks. For example, when we use ResNet50 and handcrafted features (column “Tissue ResNet”), implementation of *g* has led to an increase in ROC-AUC and PR-AUC from (0.598 ± 0.011, 0.279 ± 0.011) to (0.618 ± 0.010, 0.313 ± 0.014).

**Table 5:** Segmentation Quality-Weighted Performance. Primary performance is in ROC-AUC. Mean PR-AUC over different prediction tasks is also given in the bottom row. Comparison with unweighted predictions in Table 4 show that weighting instances by segmentation quality gives us a significant boost in performance.

| Label/Stain | Tissue<br>Resnet | Tissue<br>Resnet<br>(ATI) | Tissue<br>ResNet<br>Only | Tissue HC |
| --- | --- | --- | --- | --- |
| **ATI** | 0.652±0.021 | 0.644±0.015 | 0.654±0.012 | 0.581±0.019 |
| **DGF** | 0.591±0.022 | 0.543±0.022 | 0.569±0.022 | 0.489±0.027 |
| **DGF/SR** | 0.642±0.036 | 0.635±0.037 | 0.613±0.027 | 0.562±0.033 |
| **Remuzzi A** | 0.589±0.028 | 0.601±0.027 | 0.589±0.028 | 0.625±0.024 |
| **Remuzzi G** | 0.569±0.033 | 0.56±0.048 | 0.562±0.038 | 0.506±0.038 |
| **Remuzzi IF** | 0.648±0.025 | 0.6±0.027 | 0.589±0.021 | 0.548±0.035 |
| **Remuzzi TA** | 0.585±0.073 | 0.596±0.045 | 0.562±0.056 | 0.538±0.06 |
| **Mean**<br>**ROC-AUC** | **0.618±0.01** | **0.609±0.009** | **0.617±0.008** | **0.563±0.011** |
| **Mean**<br>**PR-AUC** | **0.313±0.014** | **0.291±0.013** | **0.292±0.012** | **0.261±0.014** |

While weighting tissues by segmentation quality has provided a performance boost for most prediction tasks, the boost is largest for the prediction of Remuzzi IF (average ROC-AUC improvement: 0.101 ± 0.023). One possible explanation is that interstitial fibrosis is a histological change not so well captured by our featuresets as the changes reside between, rather than within, the segmented tissues. Fibrosis also correlates strongly with tubular atrophy, which is characterised by changes in the basement membrane near tissue boundaries. Neither of these would be visible if objects are under-segmented. In our case, most instances with poor segmentation quality are also the ones that are undersegmented due to how the segmentation results were combined (Equation 3). Consequently, these tissues also become less relevant to the prediction task.

There may be a second reason why the proposed weighting improves predictions. As biopsies tend to contain many tissues, it is often not possible to delineate every tissue within a slide. The annotator may have the tendency to choose regions where tissues are legible and create training sets based on these examples. Even though the delineation task was not performed by an expert, the choice of tissues on its own may still contain information indicative of what constitutes as “good quality” or what counts as “relevant”.

## 4. Conclusion and Further Work

We propose a scheme to improve the performance of multi-instance prediction tasks based on soft attention models by incorporating weak labels at the local level. This approach could make projects that are currently bottle-necked by expert annotations more scalable. In our case, the weak labels are the tissues’ outlines delineated into coarse classes. These delineated tissue instances provided several advantages over featuresets extracted from rectangular tiles. Firstly, having features extracted from tissues allow us to design handcrafted features inspired by domain knowledge. We show that the generalised performance is improved significantly when handcrafted features are combined with deep features compared to models trained using only either deep or handcrafted features. Secondly, features from rectangular tiles do not generally conform to the functional boundaries of tissue compartments. Using features based on tissues in soft attention models allows an intuitive visualisation scheme pinpointing relevant tissues for transparent diagnostics. Thirdly, we argue that the attention values predicted could only be meaningful if the images resemble the training data. Instances significantly different from the training set could contribute to noise at the bag-level prediction so we propose to incorporate a tissue quality metric derived from an ensemble of BNNs to reduce their impact.

There are also limitations that need to be addressed in our experiments. Firstly, it is not clear whether the advantage of combining handcrafted features with deep features will still hold for larger datasets. Secondly, when there are multiple relevant instances in a slide, soft attention would only focus on a small number of instances in most cases. Many relevant instances are predicted low values. Therefore, it remains challenging to quantify the number of relevant tissues in a slide - this will be needed to quantify uncertainties of slide-level predictions. We will need to investigate whether it can be solved using an ensemble of models.

Thirdly, we find that there are currently insufficient diseased cases in our dataset. For example, there are currently only several dozens of sclerosed glomeruli from all datasets combined, hence the segmentation performance may not generalise well to diseased instances. In many cases, if the segmentation of diseased instances is suboptimal, soft attention may focus on tissues with confounding visual changes. This would result in correct slide-level prediction but a wrong focus of attention. Isolating highly correlated pathological changes may be possible through arithmetic operations with soft attention maps, but this may require a sufficiently large training dataset.

In some cases, we recognise that performance of multi-instance learning may still not match models trained by fully-supervised approaches. If full-supervision is needed, our proposed platform can be used as a guide for a human-in-a-loop labelling scheme to bootstrap a project. For example, we can choose to label specifically only tissue compartments with high attention from slides that have been given a wrong prediction. The soft attention model can then be repeatedly re-trained with labelled instances excluded from the slide in each iteration. This may help to reduce the time needed for pathologists to go through the entire dataset when we want to add a new diagnosis.

## Data Availability

All data produced in the present study are available upon reasonable request to the authors.

## Acknowledgement

KHT is funded by the EPSRC and MRC grant number EP/L016052/1. JR is supported by the Oxford NIHR Biomedical Research Centre and the PathLAKE consortium (Innovate UK App. Nr. 18181). JK is supported by the Dutch Kidney Foundation (Grant No. 17OKG23) and the Human(e) AI Research Priority Area by the University of Amsterdam. MK is supported by NHS Blood and Transplant and QUOD sample analysis funded by Kidney Research UK funding KS RP 002 20210111 awarded to MK. The authors thank the UK QUOD Consortium and NHS Blood and Transplant UK Registry for the clinical samples and metadata analyzed in this study.

## Footnotes

1 Collection of QUOD samples and the research ethics approval was provided by QUOD (NW/18/0187).

2 Research ethics approved by the National Ethics Review Committee of the United Kingdom (12/EE/0273)

3 Ethics approved by the Research Ethics Committee of Oxford University Hospital NHS Foundation Trust Research and Governance (19/WM/0215)

## Notes

### Competing Interest Statement

The authors have declared no competing interest.

### Author Declarations

Use of QUOD samples: NHS Health Authority, National Research Ethics Service, gave ethical approval for this work (IRAS project ID: 87824; Quality in Organ Donation (QUOD) (NW/18/0187), sponsored by University of Oxford.) Use of native biopsies: Research Ethics Committee of Oxford University Hospital - NHS Foundation Trust Research and Governance gave ethical approval for this work. (19/WM/0215) Use of NMP dataset: The National Ethics Review Committee (East of England) of the United Kingdom gave ethical approval for this work. (REC reference (12/EE/0273), IRAS project ID 106793)

### Summary of Updates

References details revised. Figures 8-10 updated.

