## Supplementary Material for "Predicting Clinical Endpoints and Visual Changes with Quality-Weighted Tissue-based Renal Histological Features"

### S1. Slide Labels

Assessment of WSIs was performed according to the standard from Remuzzi et al (Remuzzi et al., 2006). ATI was graded using the following criteria: *0 – absent; 1 – loss of brush borders/vacuolation of tubular epithelial cells; 2 – cell detachment/cellular casts; 3 – coagulation necrosis.*

The number of slides with labels available varies depending on the prediction tasks (eg. for slides that do not contain enough arteries are not scored for Remuzzi A) and is summarised in Table S1.

Table S1: **Number of Slide Labels Available**

| Label / Stain | Donor / Slides (of which QUOD Dataset) |
|---|---|
| ATI / PAS | 170/361 (135/145) |
| DGF / PAS | 283/321 (283/321) |
| DGF / SR | 143/143 (143/143) |
| Remuzzi A / PAS | 89/95 (89/95) |
| Remuzzi G / PAS | 137/163 (133/143) |
| Remuzzi IF / PAS | 170/311 (135/145) |
| Remuzzi TA / PAS | 135/145 (135/145) |

### S2. Localised Tissue Assessment

Delineation of tissues had been curated incrementally throughout the workflow's development. At earlier stages where we had fewer annotations, a UNet was trained based on a smaller training set. At this stage, a subset of objects,

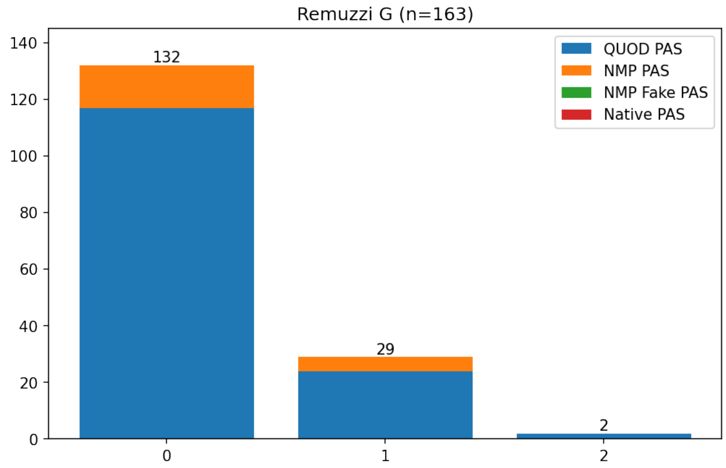


(a) Remuzzi Grade G

Figure S1: **Distribution of grades given by a renal pathologist.** Slides that do not contain enough glomeruli/vessels are not scored for that specific category. Figures S1a-S1d are standard Remuzzi Grades; Figure S1e assesses the overall acute damage in proximal tubules.

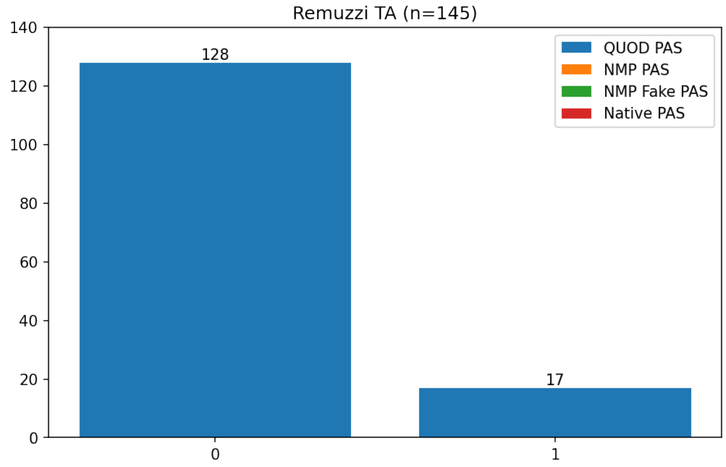


(b) Remuzzi Grade TA

Figure S1: **(cont.) Distribution of grades given by a renal pathologist.**

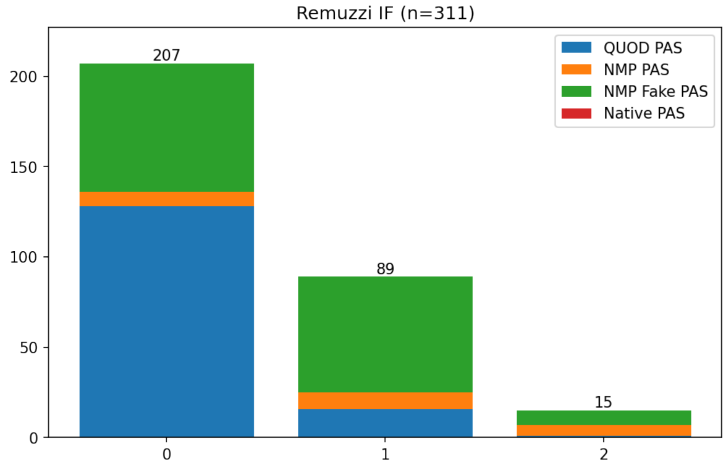


(c) Remuzzi Grade IF

Figure S1: **(cont.) Distribution of grades given by a renal pathologist.**

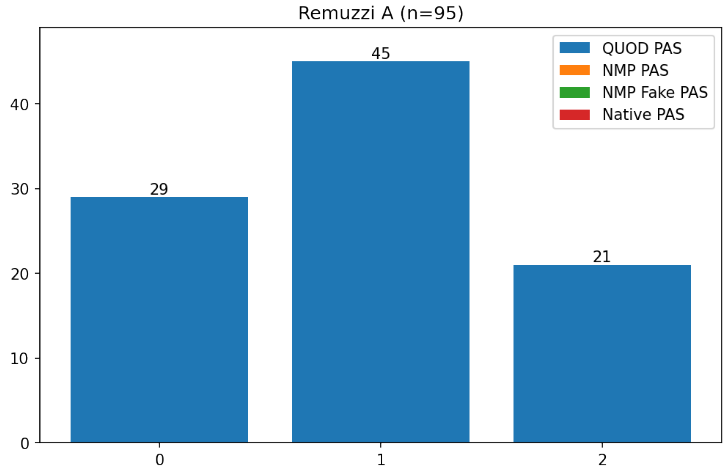


(d) Remuzzi Grade A

Figure S1: **(cont.) Distribution of grades given by a renal pathologist.**

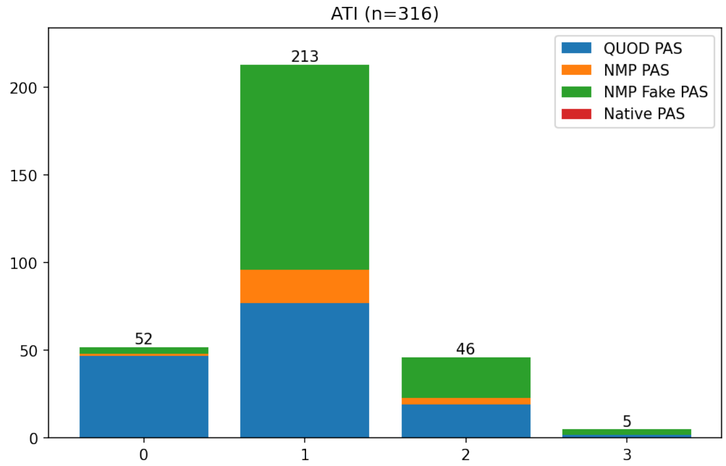


(e) ATI Grade

Figure S1: **(cont.) Distribution of grades given by a renal pathologist.**

either delineated by hand or segmented by a single UNet, has been reviewed by a renal pathologist.

This subset was originally picked by hand and assessed randomly by the pathologist. However, after assessing several dozen tissues, we narrowed down the subset further due to the pathologist's time constrain. From this point forward, the order of the tissues assessed was chosen to maximise the coverage of the tissues' Variational AutoEncoder embedding according to Sener et al. (Sener and Savarese, 2017).

These tissues are not picked at random as the assessment was intended for a different task beyond the scope of this paper. Note that the way samples were picked might have slightly exaggerated the number of artifacts in the distribution.

A total of 1992 objects had been reviewed, 1032 of which were segmented by UNets and 960 were delineated by hand. These objects have been labelled or predicted as belonging to either tubule or glomeruli class. The statistics of

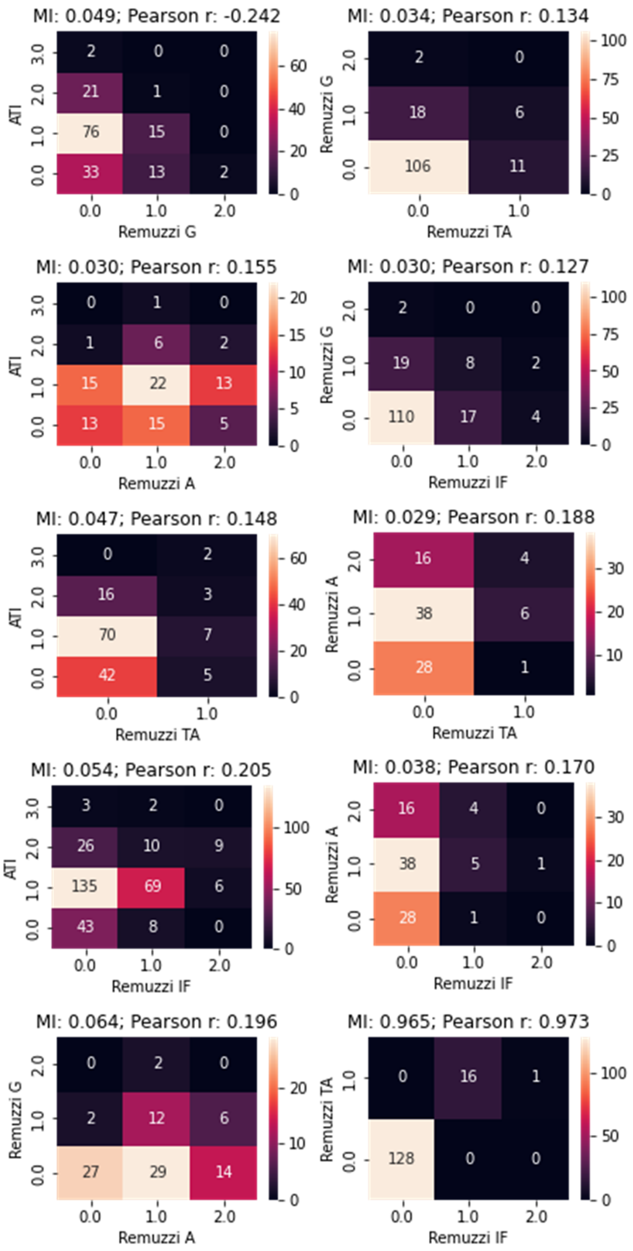


Figure S2: **Correlation between slide-level grades given by pathologist**. Normalised Mutual Information (MI) and Pearson r values are shown. Remuzzi TA and IF are highly correlated ($r = 0.973$) but we have more slides graded for IF but not TA which are not shown in the heatmap.

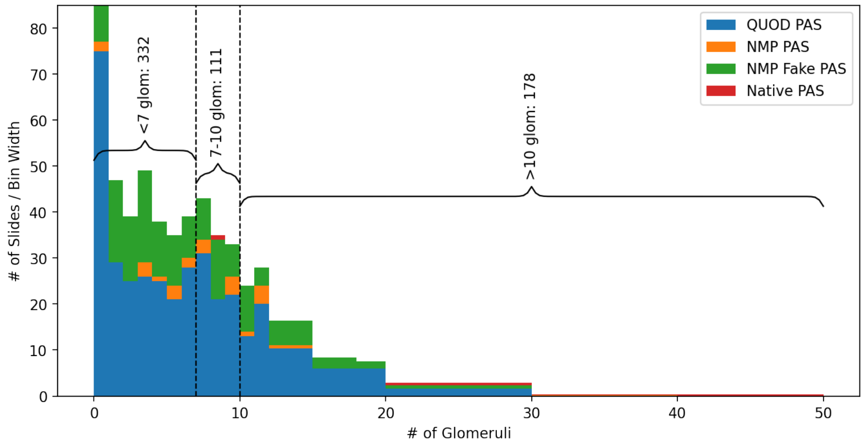


Figure S3: **Distribution of number of glomeruli in our datasets.** There is an inherent trade-off between obtaining biopsies size and risk of complications such as bleeding. Some slides contain multiple adjacent sections of the same biopsy - we manually identified these slides avoided double counting these instances. The majority (332) of slides do not have enough glomerli for assessment as stipulated by Banff Criteria.

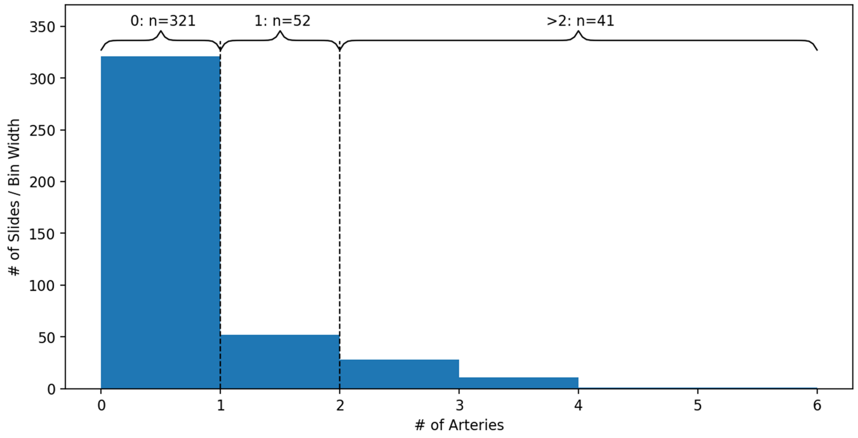


Figure S4: **Distribution of number of arteries in the QUOD dataset (PAS-stained slides only).** There are some discrepancy between the artery count and those that have received a Remuzzi A grade. This is possibly because some of the slides have a arties that are partially truncated.

| Remuzzi A/PAS AUC: row > col? | Tissue ResNet50 | Tissue ResNet50 (ATI) | Tissue VGG16 | Tissue InceptionV3 | Tissue ScatterNet | Tissue ResNet50 Only | Tissue HC | Tiles (2 Levels) ResNet50 | Tiles (2 Levels) ScatterNet | Tiles (1 Level) ResNet50 |
|---|---|---|---|---|---|---|---|---|---|---|
| Tissue ResNet50 | Und | Und | 1 | Und | 1 | 1 | Und | 1 | 1 | 1 |
| Tissue ResNet50 (ATI) | Und | Und | Und | Und | 1 | Und | Und | 1 | 1 | 1 |
| Tissue VGG16 | -1 | Und | Und | -1 | Und | Und | Und | 1 | 1 | 1 |
| Tissue InceptionV3 | Und | Und | 1 | Und | 1 | Und | Und | 1 | 1 | 1 |
| Tissue ScatterNet | -1 | -1 | Und | -1 | Und | Und | -1 | 1 | 1 | 1 |
| Tissue ResNet50 Only | -1 | Und | Und | Und | Und | Und | Und | 1 | Und | 1 |
| Tissue HC | Und | Und | Und | Und | 1 | Und | Und | 1 | 1 | 1 |
| Tiles (2 Levels) ResNet50 | -1 | -1 | -1 | -1 | -1 | -1 | -1 | Und | Und | Und |
| Tiles (2 Levels) ScatterNet | -1 | -1 | -1 | -1 | -1 | Und | -1 | Und | Und | 1 |
| Tiles (1 Level) ResNet50 | -1 | -1 | -1 | -1 | -1 | -1 | -1 | Und | -1 | Und |

Figure S5: **Comparison of AUC between different featuresets for predicting Remuzzi A.** Entries are labelled "1" or "-1" according to whether the row performs better than the column by an AUC difference greater than $\sqrt{\sigma^2_{AUC1} + \sigma^2_{AUC2}}$. $\sigma$ is the uncertainty estimate of the mean as listed in Table 3. Entries where the difference is smaller than this threshold are labelled "Und". Here we can see an overall trend where tissue features outperform those from tile features.

| mean (inv. var weighted) AUC: row > col? | | Weighted | | | | Unweighted | | | |
|---|---|---|---|---|---|---|---|---|---|
| | | Tissue Resnet50 Metadata | Tissue Resnet50 | Tissue Resnet50 Only | Tissue HC | Tissue Resnet50 Metadata | Tissue Resnet50 | Tissue Resnet50 Only | Tissue HC |
| Weighted | Tissue Resnet50 Metadata | Und | 1 | 1 | 1 | 1 | 1 | 1 | 1 |
| | Tissue Resnet50 | -1 | Und | Und | 1 | 1 | 1 | 1 | 1 |
| | Tissue Resnet50 Only | -1 | Und | Und | 1 | 1 | 1 | 1 | 1 |
| | Tissue HC | -1 | -1 | -1 | Und | -1 | -1 | 1 | 1 |
| Unweighted | Tissue Resnet50 Metadata | -1 | -1 | -1 | 1 | Und | Und | 1 | 1 |
| | Tissue Resnet50 | -1 | -1 | -1 | 1 | Und | Und | 1 | 1 |
| | Tissue Resnet50 Only | -1 | -1 | -1 | -1 | -1 | -1 | Und | Und |
| | Tissue HC | -1 | -1 | -1 | -1 | -1 | -1 | Und | Und |

Figure S6: **Comparison of Mean AUC between Featuresets Unweighted vs Weighted by Segmentation Quality**. labelling scheme is the same as Figure S5. Here we can see an overall trend where AUC is higher when weighted.

tissues are shown in Tables S2 and S3. From Table S2 it can be seen that less than half (157/340) of the tissues labelled as "Tubules" were actually proximal tubules, the rest were objects irrelevant for assessment. However, since this is an on-going project, the quality of the delineation might have improved over time. A qualitative estimate showed that approximately 70% of the delineated tubules are proximal in the up-to-date dataset.

A total of 731 proximal tubules have been reviewed by the pathologist if we include tissues that have been wrongly labelled as "glomeruli". These proximal tubules are graded for chronic (TA 0-5) and acute (ATI 0-5) damages. TA was graded according to the amount of thickening of the basement membrane: *0: absent; 1: mild thickening; 2: significant thickening but to an extent less than the thickness of epithelial cells; 3: thickening equal to the thickness of healthy epithelial cells; 4: thicker than healthy epithelial cells; 5: reserved for extreme cases.* ATI was graded as follows: *0 – absent; 1 – segmental/local loss of brush borders/vacuolation of tubular epithelial cells; 2 - total loss of brush borders/vacuolation 3 – cell detachment/cellular casts; 4 – coagulation necrosis; 5 - reserved for extreme cases.* The distribution of these grades, broken down by dataset, are shown in Figure S7. It can be seen that TA grades are heavily imbalanced. Between the 2 large datasets (QUOD and NMP), only 8 tissues have been given grade 1 and none have grades above 1. The distribution of ATI grades, on the other hand, is much more evenly spread.

Table S2: **Delineated/Segmented Tissues Reviewed by Pathologist.**

| | Segmented | Delineated |
|---|---|---|
| Tubule class | 838 | 340 |
| of which Proximal Tubules | 553 | 157 |
| Glomeruli class | 194 | 620 |
| True Positive Glomeruli | 83 | 600 |

Table S3: **Summary of Tissues Assessed by Pathologist.**

| | Segmented | Delineated |
|---|---|---|
| Proximal Tubules | 571 | 160 |
| Glomeruli | 158 | 601 |
| Vessels | 11 | 0 |
| Other Tissues | 209 | 153 |
| Artifact | 83 | 46 |
| Total Relevant | 740 | 761 |
| Total | 1032 | 960 |

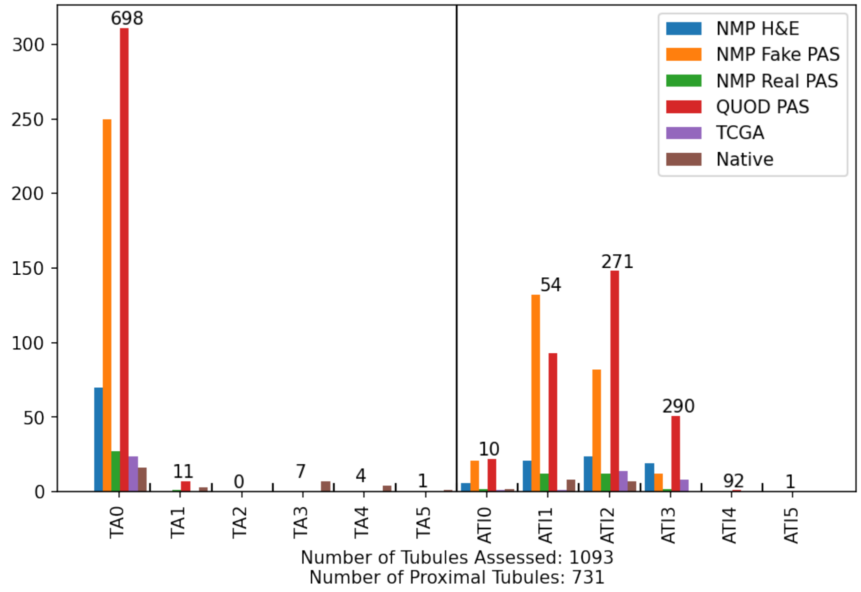


Figure S7: **Distribution of Local Grades for Proximal Tubules**

### S3. Tissue Segmentation

Details of the UNet architecture used in this study is shown in Table S4. A value of $m = 16$ is used for our default UNets (identical to Tam et al. (2020)). For the segmentation of cell nuclei, we use $m = 1$ for Block 6-8 and $m = 8$ for other blocks. For the segmentation of tissues at $1.76mpp$, we use $m = 8$ for Block 6, 8 and $m = 4$ for Block 7. A smaller number of filters is used to save memory resources as large receptive field is less relevant for the segmentation of cell nuclei and in the segmentation at low magnification. In addition to the foreground tissue classes, each UNet also outputs a background class and a boundary class. After obtaining instances using max-flow-min-cut, we expand the area of each instance by looping through each instance iteratively and dilate each mask with a 3x3 kernel until the instance exceeds the boundary or touches another instance. We find the inclusion of boundary class helps to separate tissues with ambiguous boundaries.

Ideally we would want the soft values from the UNet ensemble to scale linearly with the probabilities for correct class prediction. However, we find this is not the case for data-limited tissue classes such as glomeruli.

Figure S8 shows segmentation results on the QUOD-PAS slides. The plots show how the soft values of the combined segmentation scale with $A$, the multiplier of $\sigma$ in Equation 3. For each value of $A$, we calculate a histogram binning all pixels predicted a certain value $\tilde{p}$ by the UNet ensemble. The predicted probabilities $\tilde{p}$ is plotted against the actual probabilities in (b) and (d) for different values of $A$. Then we compute the L2 difference between the array $\tilde{p}$ against the actual probabilities $p$ as shown in (a) and (c).

It can be seen that the optimal values are $A = 0$ for tubules (Figures S8a-b) and $A = 2$ for glomeruli (Figure S8c-d). These results suggest that while outputs from the Bayesian network ensemble scale linearly with class probabilities when class labels are abundant, this linear relationship breaks down when uncertainties are data-limited and some empirical corrections might be needed.

Note that Equation 3 serves to remove areas that are overconfident but would

Table S4: **UNet architecture**

| Up / Down / Center Block $(n_1, n_2)$ | |
|---|---|
| $3 \cdot 3 \cdot n_1 \cdot n_2$ Conv., Inst. Norm, ReLU, 0.2 (Dropout) | |
| $3 \cdot 3 \cdot n_2 \cdot n_2$ Conv., Inst. Norm, ReLU, 0.2 (Dropout) | |
| $2 \times 2$ MaxPool / Bilinear Interpolation / - | |
| UNet Architecture | |
| Block 1 | Center $(3, m)$ |
| Block 2 | Down $(m, 2m)$ |
| Block 3 | Down $(2m, 4m)$ |
| Block 4 | Down $(4m, 8m)$ |
| Block 5 | Down $(8m, 16m)$ |
| Block 6 | Down $(16, 32m)$ |
| Block 7 | Down $(32m, 64m)$, Center $(64m, 64m)$ |
| Block 8 | Up $(96m, 32m)$ |
| Block 9 | Up $(48m, 16m)$ |
| Block 10 | Up $(24m, 8m)$ |
| Block 11 | Up $(12m, 4m)$ |
| Block 12 | Up $(6m, 2m)$ |
| Block 13 | Up $(3m, m)$ |
| Block 14 | Center $(m, OutputClasses)$ |

not add to under-confident areas. Thus, the final number of tissues detected is likely to be underestimated. This bias is introduced to offset the asymmetrical consequences of false-positive detection compared to a false-negative: while a false-negative may simply result in fewer tissues being processed, a false-positive detection would lead to misleading information being introduced into the workflow. The former case is far easier to deal with as we can simply flag a slide as "Needs Review" if we detect too few tissues.

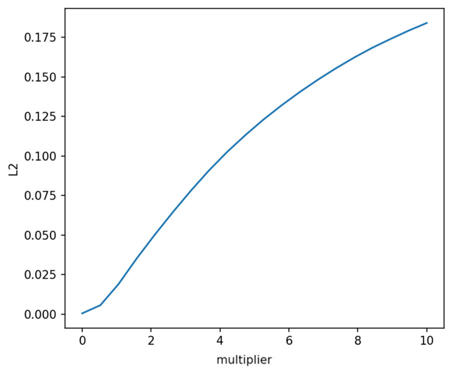


(a) L2 vs $A$ for the tubule class.

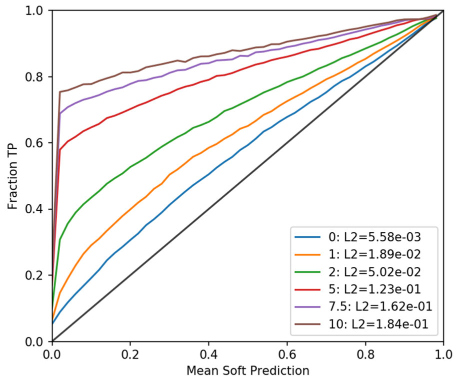


(b) Soft values for the tubules class.

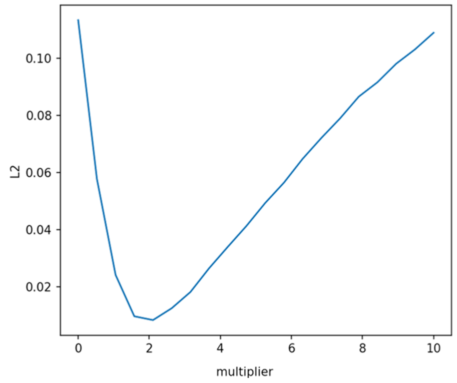


(c) L2 vs $A$ for the glomerulus class.

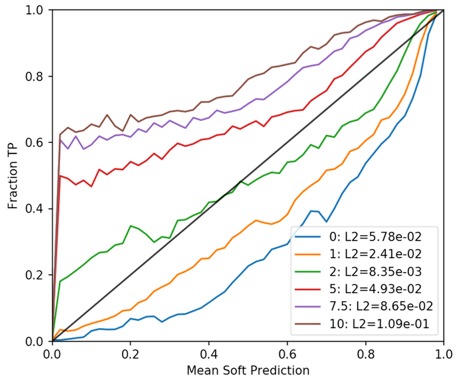


(d) Soft values for the glomerulus class.

Figure S8: **Class Probabilities vs Ensemble Averaged Segmentation Predictions**. (a) and (c) show the L2 distance between values predicted by the UNet ensemble and the actual probabilities a pixel belongs to the tubule/glomerulus class at different values of $A$. (b) and (d) show how the calibrated soft values compare with the actual probabilities a pixel belongs to a class.

### S4. Donor and Recipient Metadata

A number of metadata are available in the QUOD dataset. Based on metadata alone, the strongest predictors for the onset of DGF are donor and recipient age. These two metadata are correlated as kidneys from older donors are generally matched to older recipients due to ethical reasons. Predicting the presence of DGF based on these variables alone would give us ROC-AUC of 0.584 and 0.529 respectively. Other metadata used in the main paper are summarised in Table S5. Note that some metadata are categorical. To utilise these in neural networks, we converted them into one-hot representations. Metadata is concatenated to each tissue's feature vector, resulting in 299 extra features. Missing values and normalisation are processed in the same way as other features.

Table S5: **Description of metadata available for transplantation.**

| # | Description |
|---|---|
| 1 | Calculated Reaction Frequency at Transplant |
| 2 | Dialysis Status at Transplant |
| 3 | Donor (History of) Hypertension |
| 4 | Donor (History of) Hypotension |
| 5 | Donor Age |
| 6 | Donor Blood Group |
| 7 | Donor Blood Rhesus |
| 8 | Donor Body Mass Index |
| 9 | Donor Cause of Death |
| 10 | Donor Cytomegalovirus Test Results |
| 11 | Donor Diabetes |
| 12 | Donor Ethnicity |
| 13 | Donor Family History of Cardiovascular Disease |
| 14 | Donor Family History of Diabetes |
| 15 | Donor Gender |

Continued on next page

**Table S5 – continued from previous page**

| # | Description |
|---|---|
| 16 | Donor Gender |
| 17 | Donor Height |
| 18 | Donor Hepatitis C Virus Results |
| 19 | Donor History of Cardiovascular Disease |
| 20 | Donor History of Liver Disease |
| 21 | Donor Homozygous/Heterozygous at A Locus |
| 22 | Donor Homozygous/Heterozygous at B Locus |
| 23 | Donor Homozygous/Heterozygous at DR Locus |
| 24 | Donor Kidney Estimated Glomerulus Filtration Rate (eGFR) |
| 25 | Donor Number of Occasions with Hypotension |
| 26 | Donor Number of Occasions with Hypertension |
| 27 | Donor Type (DBD/DCD) |
| 28 | Donor Weight |
| 29 | HLA Mismatch Groups |
| 30 | Kidney Cold Ischemic Time |
| 31 | Machine Reperfusion (None/Normothermic/Hypothermic) |
| 32 | Matchability |
| 33 | Match Grade |
| 34 | Perfusate Used |
| 35 | Perfusion Quality |
| 36 | Points Score Based on Current Matchability Points Band |
| 37 | Primary Renal Disease (Categorical) |
| 38 | Recipient Age |
| 39 | Recipient Body Mass Index |
| 40 | Recipient Cytomegalovirus Test Results |
| 41 | Recipient Ethnicity |
| 42 | Recipient Gender |

Continued on next page

**Table S5 – continued from previous page**

| # | Description |
|---|---|
| 43 | Recipient Height |
| 44 | Recipient Height |
| 45 | Recipient Hepatitis C Virus Results |
| 46 | Recipient Homozygous/Heterozygous at A Locus |
| 47 | Recipient Homozygous/Heterozygous at B Locus |
| 48 | Recipient Homozygous/Heterozygous at DR Locus |
| 49 | Recipient Waiting Time |
| 50 | Status of Dialysis Prior to Transplant |
| 51 | Time Between Admission and Aorta Flushing |
| 52 | Time Between Admission and Circulatory Arrest |
| 53 | Time Between Admission and In Situ Cold Perfusion |
| 54 | Time Between Admission and Ventilation Ceased |
| 55 | Time Between Admission and Withdrawal of Support |
| 56 | Time Between Aortic Perfusion and Circulatory Arrest |
| 57 | Time Between Aortic Perfusion and Time Systolic BP to Below 50mmhg |
| 58 | Time Between Circulatory Arrest and In Situ Cold Perfusion |
| 59 | Time Between Circulatory Arrest to Retrieval for DCD Donors |
| 60 | Time Between Second Brain Stem Death to Organ Retrieval for DBD Donors |
| 61 | Total Warm Ischaemic Time |
| 62 | Whether Recipient was Highly Sensitised |

### S5. Native Biopsies

As the original datasets (QUOD and NMP) lack cases with moderate CKD, 12 cases of native biopsies were chosen to include patients with chronic changes. Cases are given a qualitative description by the pathologist: 3 cases with no

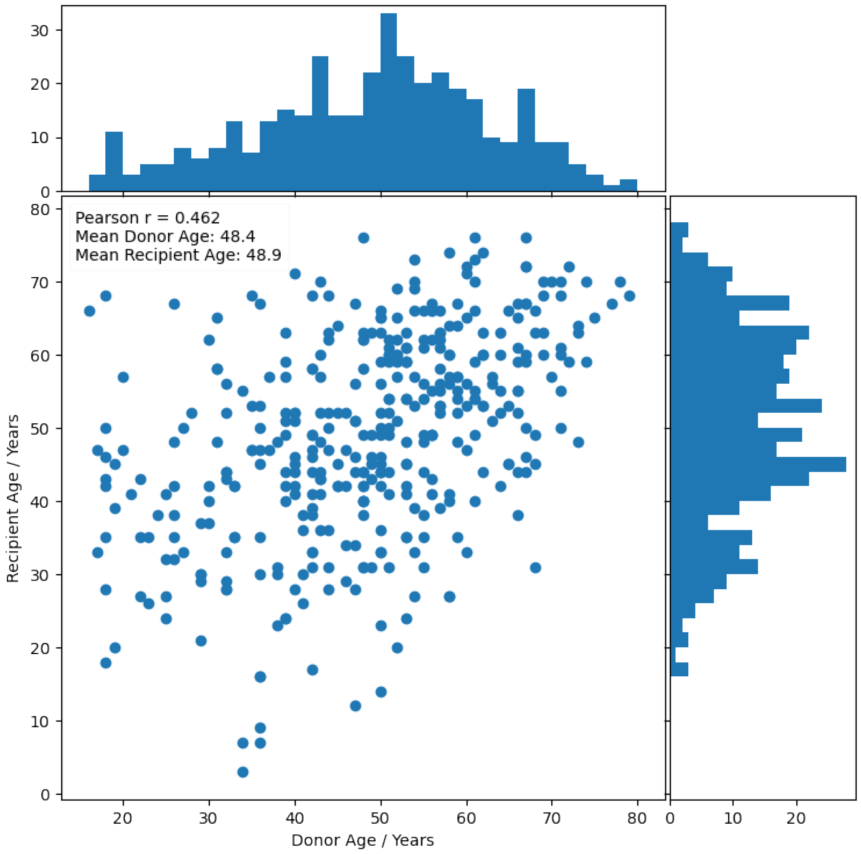


Figure S9: **Scatter plot showing how kidneys from older donors tend to be matched to older recipients.**

Table S6: **QUOD Donor Characteristics.** $n$ differs for each clinical variable as there are missing entries for some donors.

| **Parameter** | DBD | | DCD | |
|---|---|---|---|---|
| | **n** | **mean±std** | **n** | **mean±std** |
| Donor age in years | 180 | 47.5±14.4 | 164 | 49.7±14.1 |
| Donor BMI kg/m2 | 180 | 27.0±5.66 | 163 | 27.4±5.27 |
| Serum crea at admission in µmol/l | 174 | 81.2±33.7 | 159 | 74.9±27.4 |
| Serum crea at retrieval in µmol/l | 172 | 94.9±74.0 | 159 | 70.7±31.8 |
| Estimated GFR in ml/min/1.73m2 | 166 | 101±55.8 | 149 | 118±52.6 |
| Urine output last hour in ml | 166 | 99.9±91.9 | 152 | 123±123 |
| Urine output last 24 hours in ml | 113 | 3260±1870 | 107 | 2730±1620 |
| Cold ischemic time in hours | 180 | 14.8±4.63 | 161 | 13.5±4.58 |

chronic changes; 4 cases with ‘mild’ chronic tubular changes; 5 cases with ‘moderate’ chronic changes. Slides with inflammation, haemorrhage or potential drug effects are not present in these slides. All slides were scanned using a Philips IntelliSite scanner at x40 (0.25mpp). These slides were only used to train the segmentation part of the pipeline.

### S6. Handcrafted Features

A list of handcrafted features is shown in Table S7.

Table S7: **List of handcrafted features extracted from tissues.**

| # | **Name : Description** | **n** |
|---|---|---|
| 1 | area: *Area of the segmented tissue* | 1 |

Continued on next page

**Table S7 – continued from previous page**

| # | Name : Description | n |
|---|---|---|
| 2 | n_glom: *# of Glomeruli* | 1 |
| 3 | n_tub: *# of Tubules* | 1 |
| 4 | n_ves: *# of Vessels* | 1 |
| 5 | slide_area: *Area of slide in mmˆ2* | 1 |
| 6 | biopsy_area: *Total area of biopsy tissues in mmˆ2* | 1 |
| 7 | max_dist: *Maximum value of distance transform of the tissue — (max(D))* | 1 |
| 8 | nuclei_density: *Nuclei density; # of nuclei / area of tissue* | 1 |
| 9 | nuclei_moments_centre_max_dist: *Maximum distance of nuclei measured from centre of tissue* | 1 |
| 10 | nuclei_moments_centre_mean_dist: *Mean distance of nuclei measured from centre of tissue* | 1 |
| 11 | nuclei_moments_centre_min_dist: *Minimum distance of nuclei measured from centre of tissue* | 1 |
| 12 | nuclei_moments_centre_norm_max_dist: *Maximum distance of nuclei measured from centre of tissue, normalised by max(D) for each tissue* | 1 |
| 13 | nuclei_moments_centre_norm_mean_dist: *Mean distance of nuclei measured from centre of tissue, normalised by max(D) for each tissue* | 1 |
| 14 | nuclei_moments_centre_norm_min_dist: *Minimum distance of nuclei measured from centre of tissue, normalised by max(D) for each tissue* | 1 |

Continued on next page

**Table S7 – continued from previous page**

| # | Name : Description | n |
|---|---|---|
| 15 | nuclei_moments_kurtosis: *Kurtosis of nuclei distribution from edge* | 1 |
| 16 | nuclei_moments_max_dist: *Maximum distance of nuclei measured from edge of tissue* | 1 |
| 17 | nuclei_moments_mean_dist: *Mean distance of nuclei measured from edge of tissue* | 1 |
| 18 | nuclei_moments_min_dist: *Minimum distance of nuclei measured from edge of tissue* | 1 |
| 19 | nuclei_moments_norm_max_dist: *Maximum distance of nuclei measured from edge of tissue, normalised by max(D) for each tissue* | 1 |
| 20 | nuclei_moments_norm_mean_dist: *Mean distance of nuclei measured from edge of tissue, normalised by max(D) for each tissue* | 1 |
| 21 | nuclei_moments_norm_min_dist: *Minimum distance of nuclei measured from edge of tissue, normalised by max(D) for each tissue* | 1 |
| 22 | nuclei_moments_norm_variance: *Variance of nuclei distance from edge, normalised by max(D) for each tissue* | 1 |
| 23 | nuclei_moments_skewness: *Skewness of nuclei distance from edge* | 1 |
| 24 | nuclei_moments_variance: *Variance of nuclei distance from edge* | 1 |
| 25 | nuclei_nnuclei: *Number of nuclei per tissue* | 1 |
| 26 | nuclei_nuclei_area_050percentile: *Area of nuclei* | 10 |

Continued on next page

**Table S7 – continued from previous page**

| # | Name : Description | n |
|---|---|---|
| 27 | nuclei_nuclei_col_b_050percentile: *Nuclei colour pixel values, blue channel* | 10 |
| 28 | nuclei_nuclei_col_g_050percentile: *Nuclei colour pixel values, green channel* | 10 |
| 29 | nuclei_nuclei_col_r_050percentile: *Nuclei colour pixel values, red channel* | 10 |
| 30 | shape_Ixx: *sum(M_x * M_x)* | 1 |
| 31 | shape_Ixx_norm: *sum(M_x * M_x) / count(M)* | 1 |
| 32 | shape_Iyy: *sum(M_y * M_y)* | 1 |
| 33 | shape_Iyy_norm: *sum(M_y * M_y) / count(M)* | 1 |
| 34 | shape_Izz: *Moment of inertia of tissue — sum(M_x * M_x + M_y * M_y)* | 1 |
| 35 | shape_Izz_norm: *Moment of inertia of tissue, normalised by max(D). Larger value = more elongated — sum(M_x * M_x + M_y * M_y) / count(M)* | 1 |
| 36 | shape_aspect: *Minor / Major Axis ratio* | 1 |
| 37 | shape_ax1: *Major axis of tissue* | 1 |
| 38 | shape_ax1_norm: *Major axis of tissue, normalised by max(D)* | 1 |
| 39 | shape_ax2: *Minor axis of tissue* | 1 |
| 40 | shape_ax2_norm: *Minor axis of tissue, normalised by max(D)* | 1 |
| 41 | shape_convex: *Ratio of the tubule's mask over the convex hull of the mask* | 1 |
| 42 | shape_moment_mask: *np.sum(r1 * dist * mask) / np.sum(mask); r1 is radial distance from Centre of Mass of M* | 1 |

Continued on next page

**Table S7 – continued from previous page**

| # | Name : Description | n |
|---|---|---|
| 43 | shape_moment_mask_dist: *np.sum(r2 * dist * mask) / np.sum(mask); r2 is radial distance from D*M Centre of Mass of M* | 1 |
| 44 | shape_moment_mask_dist_norm: *np.sum(r2 * dist * mask) / np.sum(mask) / max(D); r2 is radial distance from D*M Centre of Mass of M* | 1 |
| 45 | tissue_bbar: *Blue-channel mean value in tissue's cytoplasm* | 1 |
| 46 | tissue_bstd: *Blue-channel std value in tissue's cytoplasm* | 1 |
| 47 | tissue_gbar: *Green-channel mean value in tissue's cytoplasm* | 1 |
| 48 | tissue_gstd: *Green-channel std value in tissue's cytoplasm* | 1 |
| 49 | tissue_rbar: *Red-channel mean value in tissue's cytoplasm* | 1 |
| 50 | tissue_rstd: *Red-channel std value in tissue's cytoplasm* | 1 |
| 51 | tissue_moment_mean_dist: *Mean distance of cytoplasm pixel values, as measured from edge of tissue — (mean((255 - img_1d[:, 0]) * distance_1d))* | 1 |
| 52 | tissue_moment_norm_mean_dist: *Mean distance of cytoplasm pixel values, as measured from edge of tissue, normalised by size of tissue — (mean((255 - img_1d[:, 0]) * distance_1d)/ max(D))* | 1 |
| 53 | tissue_moment_kurtosis: *4th order statistics of cytoplasm pixel values, as measured from edge of tissue* | 1 |
| 54 | tissue_moment_norm_variance: *Spatially-weighted (D / max(D)) variance of cytoplasm pixel values, as measured from edge of tissue, normalised by size of tissue* | 1 |

Continued on next page

**Table S7 – continued from previous page**

| # | Name : Description | n |
|---|---|---|
| 55 | tissue_moment_skewness: *Spatially-weighted (D) skewness of cytoplasm pixel values, as measured from edge of tissue, normalised by size of tissue* | 1 |
| 56 | tissue_moment_variance: *Spatially-weighted (D) variance of cytoplasm pixel values, as measured from edge of tissue, normalised by size of tissue* | 1 |
| 57 | tissue_moments_centre_mean_dist: *Mean distance of cytoplasm pixel values, as measured from centre of tissue — (255 - img_1d[:, 0]) * (max(D) - D)* | 1 |
| 58 | tissue_moments_centre_norm_mean_dist: *Mean distance of cytoplasm pixel values, as measured from centre of tissue, normalised — (255 - img_1d[:, 0]) * (max(D) - D) / (max(D)* | 1 |
| 59 | glom_bm_capsule_area: *Area of urinary space in glomerulus* | 1 |
| 60 | glom_bm_capsule_area_ratio: *(Area of Urinary Space) / (Area of Glomeruli)* | 1 |
| 61 | ves_lumen_area: *Lumen area in vessels* | 1 |
| 62 | ves_lumen_ratio: *Ratio of lumen to total area in vessels* | 1 |
| | **Total number of features** | 98 |

Table S8: **Description of the featuresets presented in this study.** Corresponds to Table 3 in the main text.

| # | Featureset | Description |
|---|---|---|
| 1 | Tissue<br>ResNet | Handcrafted combined with ResNet50 features<br>pretrained with ImageNet |

| | | |
|---|---|---|
| 2 | Tissue ResNet (ATI) | Handcrafted combined with ResNet50 features trained to classify ATI at the tissue level. |
| 3 | Tissue VGG16 | Handcrafted with VGG16 (ImageNet) features. |
| 4 | Tissue VAE | Handcrafted features with features from a Variational AutoEncoder trained on tissue patches. |
| 5 | Tissue InceptionV3 | Handcrafted with InceptionV3 (ImageNet) features. |
| 6 | Tissue ScatterNet | Handcrafted with ScatterNet features (2nd order). |
| 7 | Tiles (2 Levels) ResNet | ResNet50 (ImageNet) based on 256x256 tiles @0.44mpp and 1.76mpp. Features from concentric tiles concatenated. |
| 8 | Tiles (2 Levels) ScatterNet | ScatterNet features (2nd order) from 256x256 tiles @0.44mpp and 1.76mpp. |
| 9 | Tiles (1 Level) ResNet | ResNet50 (ImageNet) based on 256x256 tiles @0.44mpp. |
| 10 | Tiles (2 Levels) VAE | Features from the same VAE as (4) based on 256x256 tiles 0.44mpp. |
| 11 | Tissue ResNet Only | DNN features from ResNet50, pretrained with images from ImageNet. |
| 12 | Tissue HC | Handcrafted features only. |
| 13 | Tissue ResNet Metadata | ResNet (ImageNet) appended with clinical metadata. Categorical metadata are cast into one-hot format. |
| 14 | Tissue ResNet CLAM | Same featureset as #1 but uses CLAM model. |
| 15 | Tissue ResNet MIL | Same featureset as #1 but uses MIL model instead of soft attention. |
